# Healthcare-seeking behavior and hidden influenza-like-illness across the COVID-19 pandemic: a multi-country participatory surveillance study

**DOI:** 10.1101/2025.10.07.25337476

**Authors:** Mattia Mazzoli, Nicolò Gozzi, Lisa Hermans, Niel Hens, Gesa Carstens, Albert Jan van Hoek, Audrey Le Hegaret, Caroline Guerrisi, Clément Turbelin, Marion Debin, Vittoria Colizza, Chinelo Obi, Conall H Watson, Rebecca E Green, Adeline Dugerdil, Antoine Flahault, Daniela Paolotti

## Abstract

Assessing healthcare seeking behavior is critical to estimate Influenza-like-illness (ILI) burden, i.e., the fraction of population infected, and inform vaccination campaigns for Influenza, SARS-CoV-2 and RSV, aimed at reducing disease circulation and severe illness. The Covid-19 pandemic put national healthcare systems under stress, affecting surveillance and healthcare seeking behavior of patients across demographic groups and countries. The under-reporting of influenza sentinel surveillance due to patients not consulting healthcare centers hinders consistent influenza burden estimates and hampers the understanding of influenza transmission at the international level.

We leveraged an online participatory surveillance platform, InfluenzaNet, collecting data in six European countries, to study how the pandemic impacted individual healthcare seeking behavior in ILI patients and to estimate age-specific burden of ILI cases in terms of seasonal attack rates in the digital cohorts, hence including individuals who did not visit healthcare services.

We studied drivers of healthcare seeking behavior such as demographics, socio-economic traits, comorbidities, duration and severity of symptoms, education and work status via a logistic regression model adjusted by age and sex of national populations, correcting for sampling biases.

The presence of comorbidities, the duration and severity of the symptoms, and the self-assessed cause of symptoms, reflecting patients’ self-diagnosis, were drivers for health-seeking behavior consistently found across most countries, before and during the COVID-19 pandemic.

The pandemic instead had a country-specific impact on age, gender, education and labor as determinants of healthcare seeking behavior throughout the crisis.

We estimated ILI burden across countries using a standardized case definition, and assessed the burden of ILI that went undetected due to individuals not seeking healthcare from the general practitioner (GP, family physician) or other services.

Our work highlights the advantage of integrating traditional systems with online participatory surveillance, which captures healthcare seeking behavior and helps to better assess the impact of pandemics on respiratory illnesses circulation. Further adoption of these tools has the potential to favor EU-level respiratory diseases surveillance, vaccination guidelines for Influenza, RSV and SARS-CoV-2 and a deeper understanding and management of ILI epidemiology and impact.

**Research in context:** *Evidence before this study:* Before the SARS-CoV-2 pandemic, age, gender, education, the self-assessed cause of symptoms, their severity and duration were identified as known predictors of healthcare seeking behavior due to influenza-like-illness. However, most studies on population attendance to medical service focused on single countries, without assessing the robustness of these predictors across countries. Furthermore, it is not known what the impact of the pandemic was on their significance. Assessing healthcare seeking behavior is critical to estimate the burden of influenza-like-illness and inform decisions on the roll out of vaccination campaigns. We performed a literature search on Google Scholar using the keywords “ healthcare seeking behavior”, “ILI burden estimate”, “influenza-like-illness burden”, “burden estimate”, “ILI burden”, with no time limits, in the English language.

*Added value of this study:* Our study highlights the value of cross-country analyses by showing that not all known predictors of healthcare seeking behavior were robust across countries. Self-assessed cause of symptoms, their severity and duration proved robust across countries and pre-pandemic and pandemic periods except where small sample sizes prevented detection of significant effects. On the other hand, the pandemic had country-specific impact on socio-demographic predictors like age, gender, education and labor. Our work highlights substantial underdetection of ILI cases with respect to those estimated by national sentinel surveillance in all countries.

*Implications of all the available evidence:* This work suggests that the pandemic did not alter healthcare seeking behavior known predictors related to symptoms duration, severity and self-assessed cause, while we observed country-specific effects on age, gender and education factors. Digital cohorts may play a decisive role in integrating surveillance systems across countries for the estimation of ILI burden including medically unattended patients.

## Introduction

Influenza-like-illness (ILI) refers to a syndrome including general and respiratory symptoms^1^, resulting from several causes including influenza viruses, rhinoviruses, adenoviruses, coronaviruses and bacteria, e.g. *Legionella*. Viruses associated with ILI can cause severe illness, such as hospitalization or death, particularly children, elders, pregnant women and individuals with comorbidities^2^. In Europe, ILI surveillance is commonly carried out by national sentinel networks of general practitioners and hospital wards, primary and secondary care units and this monitoring is necessary to estimate Influenza burden in the population^3,4^, as for example in the Netherlands where a fraction of ILI patients are tested to investigate cause of symptoms. These surveillance systems are critical to inform vaccination campaigns and help reduce hospitalization and mortality, hence mitigating healthcare system stress during influenza seasons.

Traditional approaches rely on information collected through patients accessing healthcare providers to seek assistance for their symptoms.

Unfortunately, heterogeneities in healthcare seeking behavior across demographic groups can play a major role in hampering a correct estimation of ILI burden in the population^5^.

Many factors are associated with ILI patients not consulting healthcare services, representing barriers to ILI detection from sentinel surveillance, hence distorting ILI burden estimates^6^. Several studies already identified the most recurrent drivers of healthcare seeking behavior for several diseases in multiple countries, such as disease severity, age and gender^5,7–9^, but also comorbidities, education level^6,7,10^, and psychological aspects like perceived susceptibility^6^ and perceived severity of the disease. However, whether these drivers are consistent across countries, presenting different cultural, socio-economic and labor structure background, or country-dependent, e.g. due to different social security systems^6^, and how these might have changed during the Covid-19 pandemic, are questions that have been so far overlooked.

Previous works analyzed healthcare seeking behavior by relying on telephone^11^ or in-person surveys^5,10,12,13^, insurance data^14^, consumer platforms^15^ and one-shot web-based interviews^16^. Classic surveys are costly, often deployed *una-tantum* in single communities and structured to collect information for a specific research question. This hampers the possibility of re-using the surveys for further studies, observing temporal trends and collecting information in a consistent way across multiple communities and countries. In addition, the context may rapidly change, as during pandemics, when stay at home orders alter consultation rates and detection of circulating diseases competes with laboratories’ capacity for SAR-CoV-2 detection. Digital cohorts, instead, allow us to study the evolution in time, at high resolution, of socio-demographic traits, together with epidemiological and psychological aspects associated with health-related behaviors at high temporal resolution, such as healthcare seeking behavior or vaccination hesitancy^17^. Further participatory surveillance systems, like FluNearYou (United States)^18^ or FluTracking (New Zealand)^19^, represent critical tools to enhance behavioral analyses and ILI burden estimations at the international levels.

This is why the pandemic impact on ILI surveillance needs to be clarified from a broad and cross-country perspective, both in terms of behavioral change with respect to medical attendance in the pre-pandemic period and in terms of burden of disease that went undetected from national surveillance systems. Here we rely on InfluenzaNet^20^, a multi-country online participatory surveillance system based on a voluntary weekly survey focused on tracking symptoms of respiratory diseases and participants’ behavior, in a coherent way across countries^21^. We analyze participants’ ILI symptoms and their behavior towards healthcare seeking in Italy, the United Kingdom, Netherlands, France, Belgium and Switzerland. Based on our results, we estimate national ILI burden from online cohorts, which includes patients that do not consult healthcare services for their symptoms, hence including burden that goes usually undetected by national surveillance systems.

Our work has practical implications for public health policies at the European level since it promotes a harmonized platform for ILI standardized surveillance and comparison of healthcare seeking behavior across countries, integrating traditional surveillance and opening the doors to EU-level response to respiratory diseases. The extension of the adoption of online participatory surveillance systems may come in help of countries without well designed surveillance networks.

## Results

### Descriptive statistics and explanatory variables

We collected data on InfluenzaNet participants’ symptoms and behaviors during influenza seasons since 2011 for Italy and France, since 2016 for the UK and Switzerland, since 2020 for the Netherlands and since 2021 for Belgium. We focused on the survey responses reporting symptoms that are compatible with the case definition of ILI^1^ provided by the European Centre for Disease Prevention and Control (ECDC): sudden onset of symptoms, at least one symptom among fever (or feverishness), malaise, headache, myalgia and at least one among cough, sore throat and shortness of breath. We selected key explanatory variables observed as significant for healthcare seeking behavior from previous studies^6,7^, upon availability in the survey. In **Table 1** we show the occurrence of ILI episodes observed in our datasets stratified by duration and severity of symptoms and by patients’ characteristics in the six countries under study (see Methods subsection *Target and control variables* for details on categories aggregation). The dataset of ILI episodes is characterized by a high reporting from individuals between 15 and 64 years old, female and with a higher education level. To correct the sampling bias, we applied post-stratification to the digital cohorts by adjusting by age and sex of the respective national populations (see Methods subsection *Survey population weighting* for details on the weights computation). At a first glance, the overall share of healthcare seeking behavior among ILI patients varies substantially across countries, from 16.2% in the Netherlands, 17.1% in Switzerland and 20.4% in the UK, to 41.3%, 42.5% and 46.6% in Italy, Belgium and France respectively. See Supplementary **Table S1-S2** for the descriptive statistics of pre-pandemic and pandemic periods.

**Table 1.**
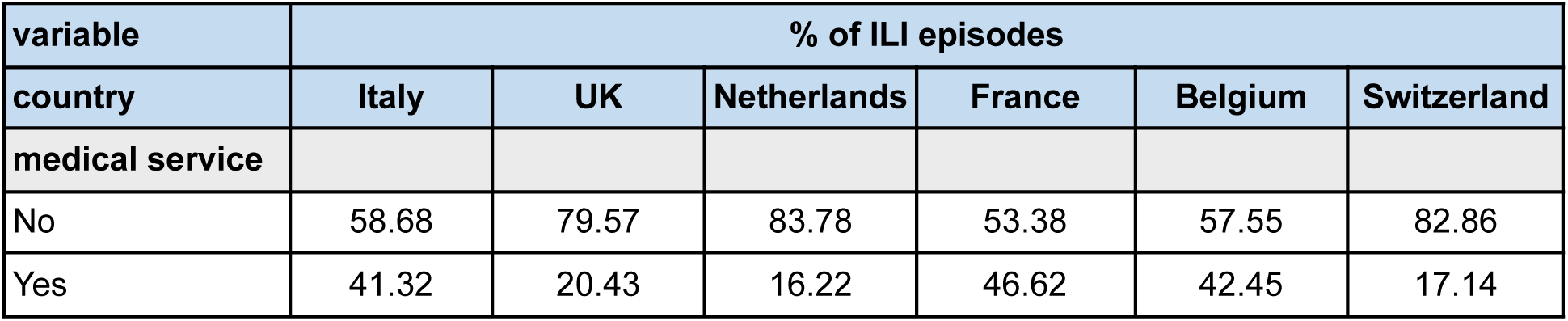
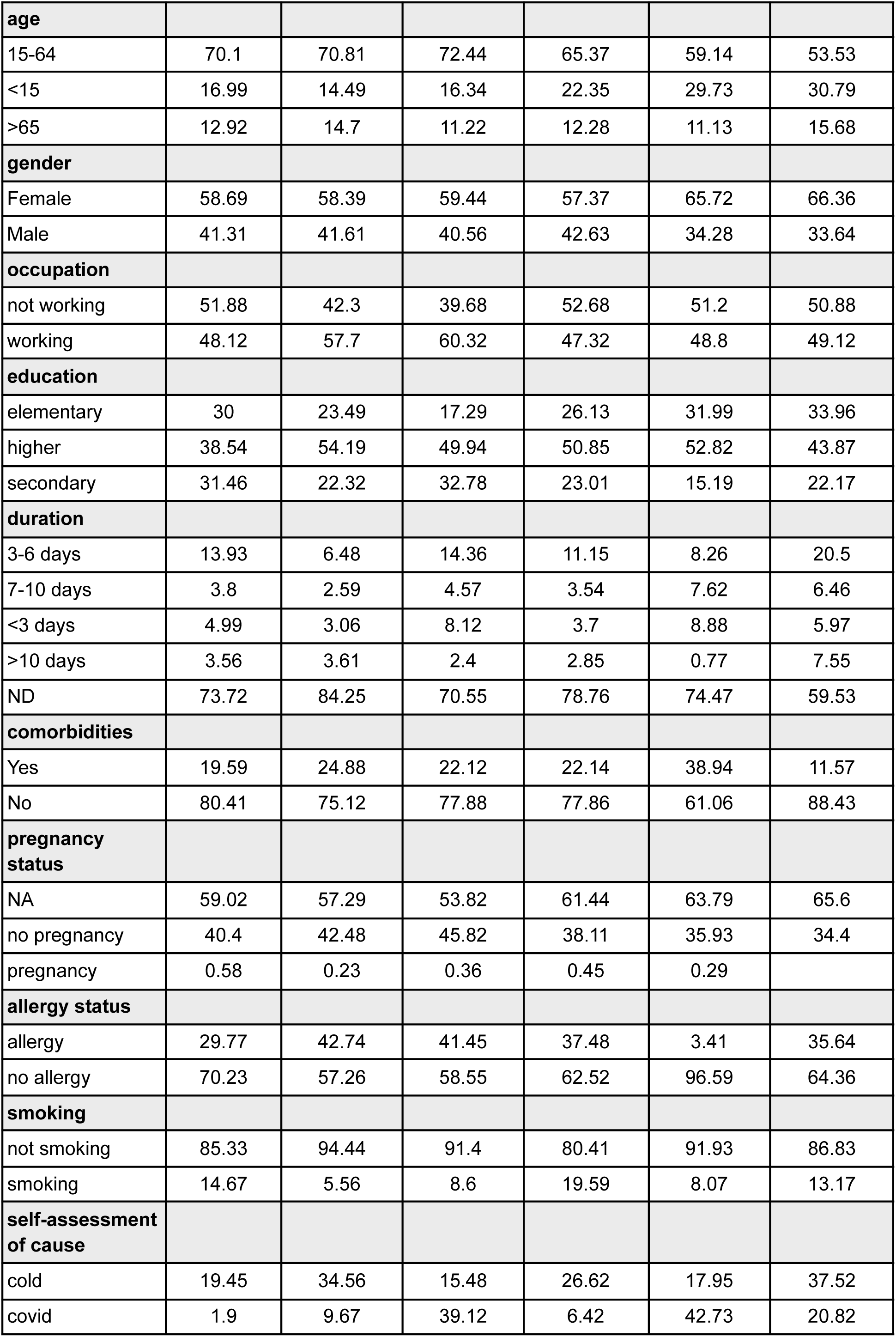
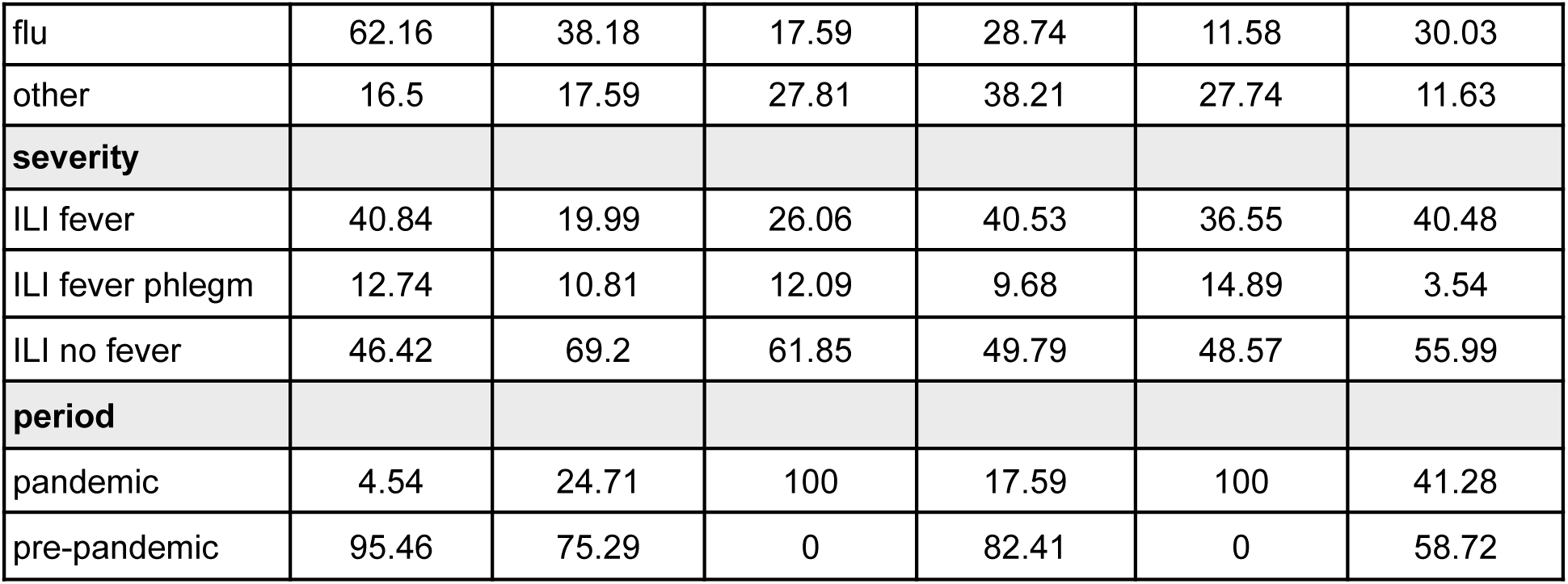
Descriptive statistics. Frequency of ILI episodes reported by InfluenzaNet individuals in Italy, the UK, the Netherlands, France, Belgium and Switzerland adjusted by age and sex of respondents. Table shows for each country and category the percentage of ILI episodes that occurred in the pre-pandemic and pandemic period. Individuals account for multiple ILI episodes across years. See Supplementary Tables S2-S3 for descriptive statistics specific to pre-pandemic and pandemic periods separately and Table S1 for unadjusted statistics.

| variable | % of ILI episodes |  |  |  |  |  |
| --- | --- | --- | --- | --- | --- | --- |
| country | Italy | UK | Netherlands | France | Belgium | Switzerland |
| medical service |  |  |  |  |  |  |
| No | 58.68 | 79.57 | 83.78 | 53.38 | 57.55 | 82.86 |
| Yes | 41.32 | 20.43 | 16.22 | 46.62 | 42.45 | 17.14 |
| **age** |  |  |  |  |  |  |
| 15-64 | 70.1 | 70.81 | 72.44 | 65.37 | 59.14 | 53.53 |
| <15 | 16.99 | 14.49 | 16.34 | 22.35 | 29.73 | 30.79 |
| >65 | 12.92 | 14.7 | 11.22 | 12.28 | 11.13 | 15.68 |
| **gender** |  |  |  |  |  |  |
| Female | 58.69 | 58.39 | 59.44 | 57.37 | 65.72 | 66.36 |
| Male | 41.31 | 41.61 | 40.56 | 42.63 | 34.28 | 33.64 |
| **occupation** |  |  |  |  |  |  |
| not working | 51.88 | 42.3 | 39.68 | 52.68 | 51.2 | 50.88 |
| working | 48.12 | 57.7 | 60.32 | 47.32 | 48.8 | 49.12 |
| **education** |  |  |  |  |  |  |
| elementary | 30 | 23.49 | 17.29 | 26.13 | 31.99 | 33.96 |
| higher | 38.54 | 54.19 | 49.94 | 50.85 | 52.82 | 43.87 |
| secondary | 31.46 | 22.32 | 32.78 | 23.01 | 15.19 | 22.17 |
| **duration** |  |  |  |  |  |  |
| 3-6 days | 13.93 | 6.48 | 14.36 | 11.15 | 8.26 | 20.5 |
| 7-10 days | 3.8 | 2.59 | 4.57 | 3.54 | 7.62 | 6.46 |
| <3 days | 4.99 | 3.06 | 8.12 | 3.7 | 8.88 | 5.97 |
| >10 days | 3.56 | 3.61 | 2.4 | 2.85 | 0.77 | 7.55 |
| ND | 73.72 | 84.25 | 70.55 | 78.76 | 74.47 | 59.53 |
| **comorbidities** |  |  |  |  |  |  |
| Yes | 19.59 | 24.88 | 22.12 | 22.14 | 38.94 | 11.57 |
| No | 80.41 | 75.12 | 77.88 | 77.86 | 61.06 | 88.43 |
| **pregnancy status** |  |  |  |  |  |  |
| NA | 59.02 | 57.29 | 53.82 | 61.44 | 63.79 | 65.6 |
| no pregnancy | 40.4 | 42.48 | 45.82 | 38.11 | 35.93 | 34.4 |
| pregnancy | 0.58 | 0.23 | 0.36 | 0.45 | 0.29 |  |
| **allergy status** |  |  |  |  |  |  |
| allergy | 29.77 | 42.74 | 41.45 | 37.48 | 3.41 | 35.64 |
| no allergy | 70.23 | 57.26 | 58.55 | 62.52 | 96.59 | 64.36 |
| **smoking** |  |  |  |  |  |  |
| not smoking | 85.33 | 94.44 | 91.4 | 80.41 | 91.93 | 86.83 |
| smoking | 14.67 | 5.56 | 8.6 | 19.59 | 8.07 | 13.17 |
| **self-assessment of cause** |  |  |  |  |  |  |
| cold | 19.45 | 34.56 | 15.48 | 26.62 | 17.95 | 37.52 |
| covid | 1.9 | 9.67 | 39.12 | 6.42 | 42.73 | 20.82 |
| flu | 62.16 | 38.18 | 17.59 | 28.74 | 11.58 | 30.03 |
| other | 16.5 | 17.59 | 27.81 | 38.21 | 27.74 | 11.63 |
| **severity** |  |  |  |  |  |  |
| ILI fever | 40.84 | 19.99 | 26.06 | 40.53 | 36.55 | 40.48 |
| ILI fever phlegm | 12.74 | 10.81 | 12.09 | 9.68 | 14.89 | 3.54 |
| ILI no fever | 46.42 | 69.2 | 61.85 | 49.79 | 48.57 | 55.99 |
| **period** |  |  |  |  |  |  |
| pandemic | 4.54 | 24.71 | 100 | 17.59 | 100 | 41.28 |
| pre-pandemic | 95.46 | 75.29 | 0 | 82.41 | 0 | 58.72 |

In **Figure 1**, we show trends of healthcare seeking behavior of ILI patients with respect to symptoms severity^7^. In all countries and across all seasons we found good agreement between severity of symptoms and healthcare seeking behavior, with trends for ARI and ILI without fever strongly overlapping in all countries except Switzerland. Since Acute Respiratory Infection only requires a single respiratory symptom, following previous work^7^ we classified the spectrum of severity in growing order with ARI as the least severe syndrome, followed by ILI with no fever, ILI with fever and finally ILI with fever and phlegm as the most severe syndrome^7^. We can observe an increase of HCSB for ILI with fever and ILI with fever and phlegm during the pandemic period, specifically in the seasons of 2019-2020 and 2020-2021 in Italy, in the season of 2019-2020 in the UK and the season of 2020-2021 in France, where data is available for both pre- and pandemic periods. However, seasonal increases of HCSB may be driven by other factors, such as participants’ medical conditions, like comorbidities and allergies, or other demographics like age and gender. In order to correct for this, we employ a logistic regression to adjust for patients’ characteristics reported in **Table 1** and check for changes in the population HCSB between pre-pandemic and pandemic periods.

**Figure 1.**
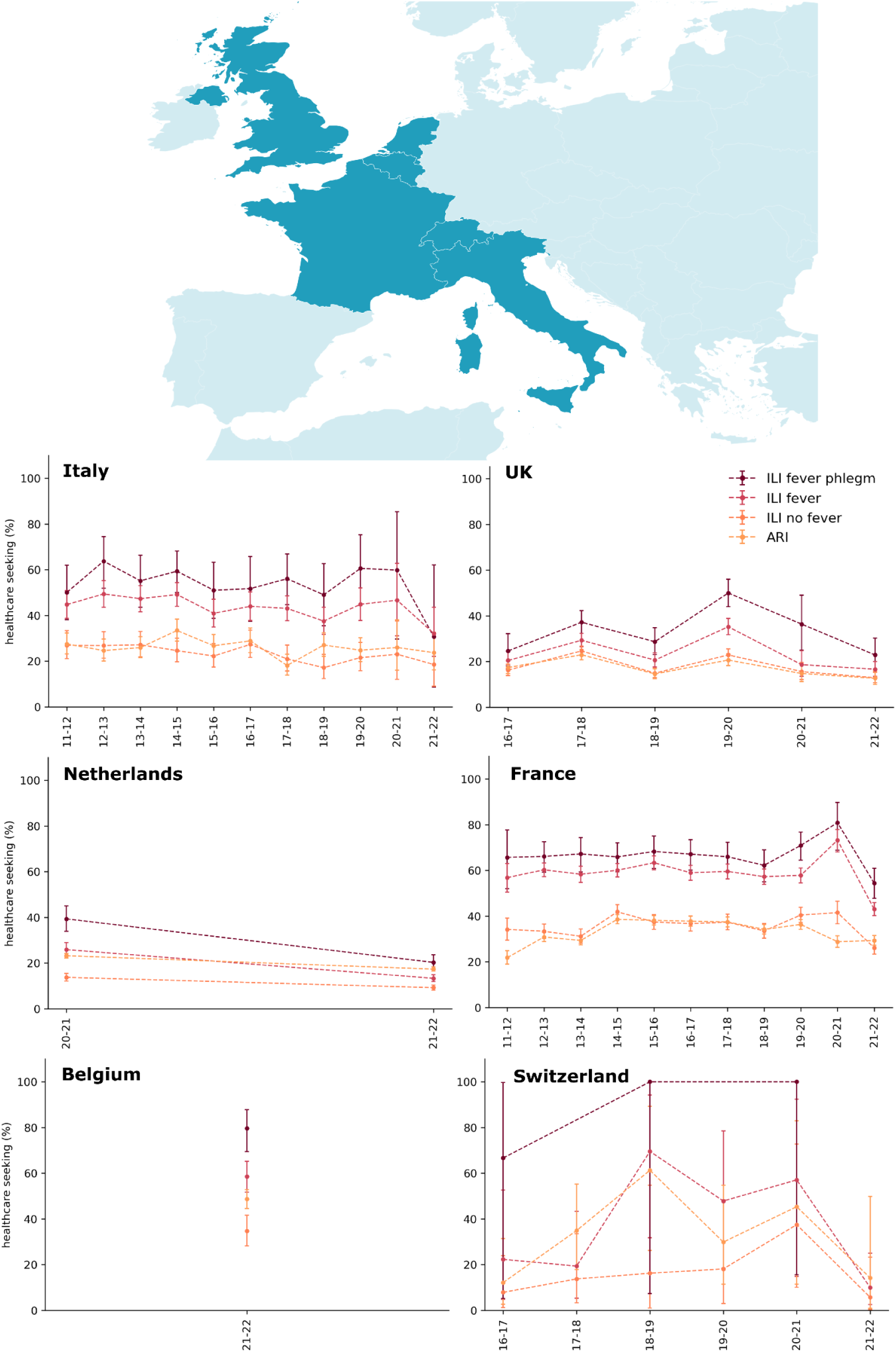
Healthcare seeking behavior by severity of symptoms. On top, a map of the countries under study. On the bottom, national comparison of adjusted healthcare seeking behavior among ILI patients from online cohorts in the six countries under study. Multiple ILI episodes from the same participants are merged together. Rates are adjusted by age and sex of participants. Darker colors represent more severe symptoms, clearer colors represent less severe symptoms. ARI (Acute Respiratory Infection) follows the ECDC case definition^1^. Whiskers represent the 95% confidence interval computed through the Clopper-Pearson method^22^. See Supplementary Figure S1 for rates treating multiple ILI episodes per participant as independent.

### Determinants of healthcare seeking behavior in the pre-pandemic and pandemic period

We quantified the impact of the Covid-19 pandemic on healthcare seeking behavior of individuals reporting ILI symptoms. As a first step, we considered only surveys submitted prior to the Covid-19 period and we performed a weighted logistic regression of the healthcare seeking behavior with respect to patients’ features reported in **Table 1**. We defined the pre-pandemic period as all the available influenza seasons prior to the 2019-2020 season. In **Table 2** we report the estimated odds-ratios and the respective confidence intervals for each explanatory variable. Odds-ratios were adjusted to account for the high incidence of the target variable (see Methods subsection *Zhang-Yu correction*). We performed this analysis on four countries over six, since InfluenzaNet was deployed in the Netherlands and Belgium only in the pandemic period.

**Table 2.**
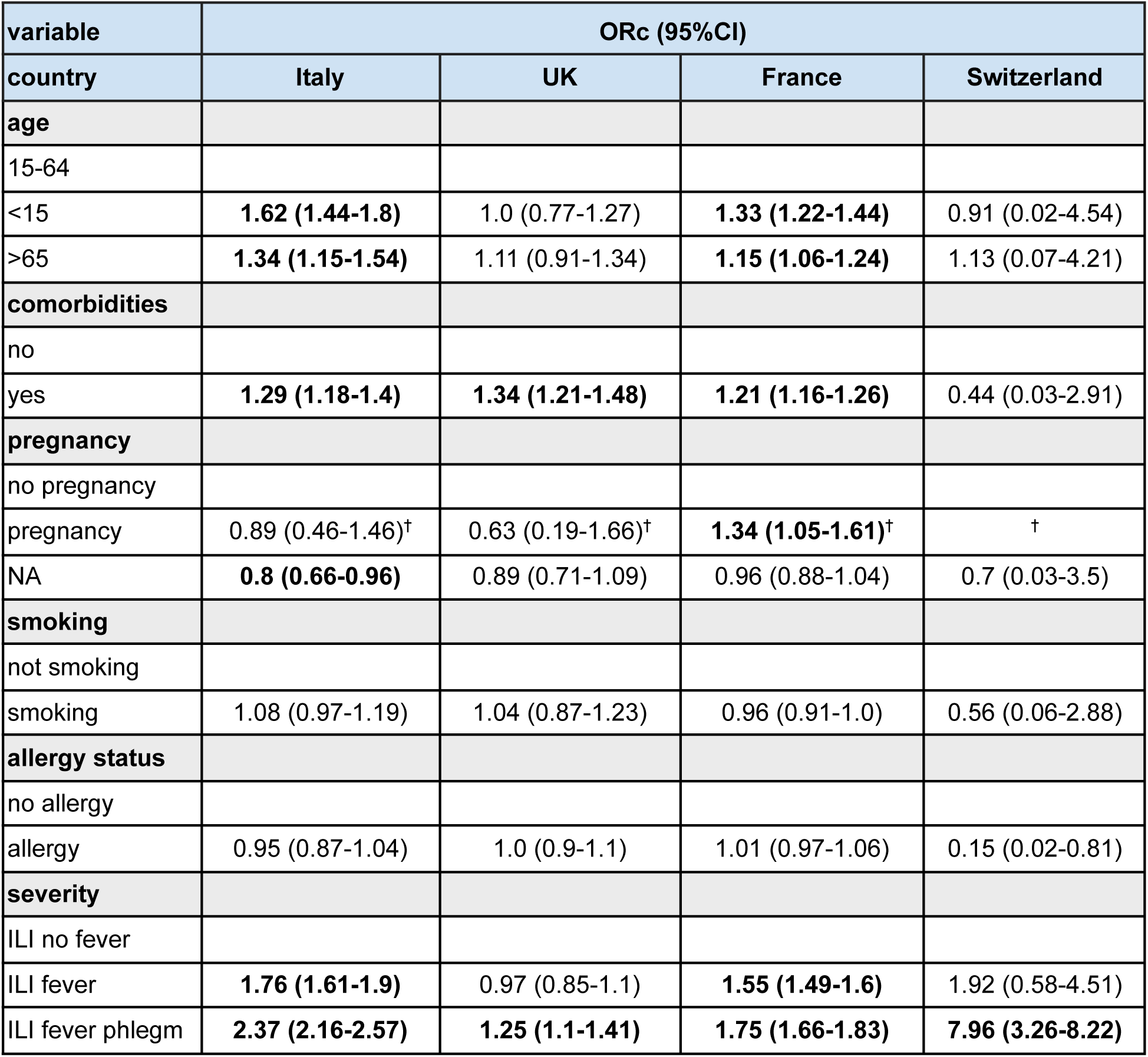
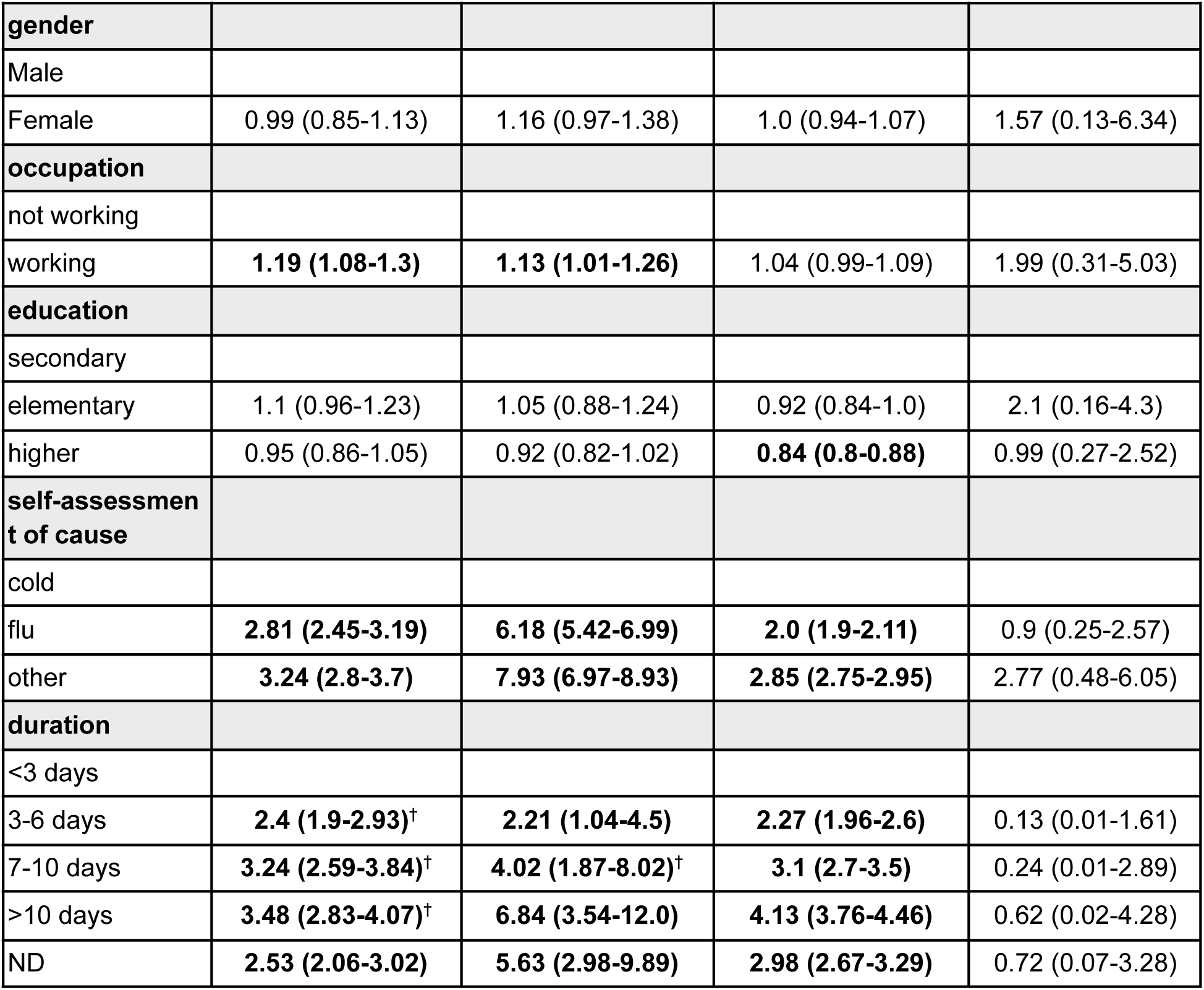
Pre-pandemic determinants of HCSB. Table shows for each country and category the adjusted odds-ratios and the 95% confidence intervals in parentheses. Reference groups for each covariate are reported in the first row. Only four countries under study collected data during the pre-pandemic period, namely Italy, the UK, France and Switzerland. Odds-ratios are adjusted by the Zhang-Yu correction. Odds-ratios for some groups are not computed due to absence of case occurrence in the class. ^†^Group sample lower than 3%. ND = No Data. Bold values refer to significant results.

| variable | ORc (95%CI) |  |  |  |
| --- | --- | --- | --- | --- |
| country | Italy | UK | France | Switzerland |
| **age** |  |  |  |  |
| 15-64 |  |  |  |  |
| <15 | **1.62 (1.44-1.8)** | 1.0 (0.77-1.27) | **1.33 (1.22-1.44)** | 0.91 (0.02-4.54) |
| >65 | **1.34 (1.15-1.54)** | 1.11 (0.91-1.34) | **1.15 (1.06-1.24)** | 1.13 (0.07-4.21) |
| **comorbidities** |  |  |  |  |
| no |  |  |  |  |
| yes | **1.29 (1.18-1.4)** | **1.34 (1.21-1.48)** | **1.21 (1.16-1.26)** | 0.44 (0.03-2.91) |
| **pregnancy** |  |  |  |  |
| no pregnancy |  |  |  |  |
| pregnancy | 0.89 (0.46-1.46)<sup>†</sup> | 0.63 (0.19-1.66)<sup>†</sup> | **1.34 (1.05-1.61)<sup>†</sup>** | <sup>†</sup> |
| NA | **0.8 (0.66-0.96)** | 0.89 (0.71-1.09) | 0.96 (0.88-1.04) | 0.7 (0.03-3.5) |
| **smoking** |  |  |  |  |
| not smoking |  |  |  |  |
| smoking | 1.08 (0.97-1.19) | 1.04 (0.87-1.23) | 0.96 (0.91-1.0) | 0.56 (0.06-2.88) |
| **allergy status** |  |  |  |  |
| no allergy |  |  |  |  |
| allergy | 0.95 (0.87-1.04) | 1.0 (0.9-1.1) | 1.01 (0.97-1.06) | 0.15 (0.02-0.81) |
| **severity** |  |  |  |  |
| ILI no fever |  |  |  |  |
| ILI fever | **1.76 (1.61-1.9)** | 0.97 (0.85-1.1) | **1.55 (1.49-1.6)** | 1.92 (0.58-4.51) |
| ILI fever phlegm | **2.37 (2.16-2.57)** | **1.25 (1.1-1.41)** | **1.75 (1.66-1.83)** | **7.96 (3.26-8.22)** |

**Table 2. Pre-pandemic determinants of HCSB.**
| **gender** |  |  |  |  |
| --- | --- | --- | --- | --- |
| Male |  |  |  |  |
| Female | 0.99 (0.85-1.13) | 1.16 (0.97-1.38) | 1.0 (0.94-1.07) | 1.57 (0.13-6.34) |
| **occupation** |  |  |  |  |
| not working |  |  |  |  |
| working | **1.19 (1.08-1.3)** | **1.13 (1.01-1.26)** | 1.04 (0.99-1.09) | 1.99 (0.31-5.03) |
| **education** |  |  |  |  |
| secondary |  |  |  |  |
| elementary | 1.1 (0.96-1.23) | 1.05 (0.88-1.24) | 0.92 (0.84-1.0) | 2.1 (0.16-4.3) |
| higher | 0.95 (0.86-1.05) | 0.92 (0.82-1.02) | **0.84 (0.8-0.88)** | 0.99 (0.27-2.52) |
| **self-assessment of cause** |  |  |  |  |
| cold |  |  |  |  |
| flu | **2.81 (2.45-3.19)** | **6.18 (5.42-6.99)** | **2.0 (1.9-2.11)** | 0.9 (0.25-2.57) |
| other | **3.24 (2.8-3.7)** | **7.93 (6.97-8.93)** | **2.85 (2.75-2.95)** | 2.77 (0.48-6.05) |
| **duration** |  |  |  |  |
| <3 days |  |  |  |  |
| 3-6 days | **2.4 (1.9-2.93)<sup>†</sup>** | **2.21 (1.04-4.5)** | **2.27 (1.96-2.6)** | 0.13 (0.01-1.61) |
| 7-10 days | **3.24 (2.59-3.84)<sup>†</sup>** | **4.02 (1.87-8.02)<sup>†</sup>** | **3.1 (2.7-3.5)** | 0.24 (0.01-2.89) |
| >10 days | **3.48 (2.83-4.07)<sup>†</sup>** | **6.84 (3.54-12.0)** | **4.13 (3.76-4.46)** | 0.62 (0.02-4.28) |
| ND | **2.53 (2.06-3.02)** | **5.63 (2.98-9.89)** | **2.98 (2.67-3.29)** | 0.72 (0.07-3.28) |

We found significant associations of higher healthcare seeking behavior with the presence of comorbidities, flu and other self-assessed causes of symptoms with respect to cold, with longer duration and higher severity of symptoms, which were consistent across countries.

For Switzerland we did not find significant factors of healthcare seeking behavior beyond symptoms severity, mostly due to pre-pandemic scarce samples, hence no conclusive results can be derived.

We found country specific associations of medical service seeking for ILI patients: age is a significant factor in Italy and France, where individuals younger than 15 and older than 65 years old are those who most sought medical attention; pregnancy is a significant factor in France and working status is a significant factor of higher healthcare seeking in Italy and the UK.

We repeated the analysis presented in the previous section considering the surveys submitted in the pandemic period, which in our dataset includes seasons of 2020-2021 and 2021-2022 for all six countries. We excluded the season of 2019-2020 as it represented a transition season. We report the estimated odds-ratios and corresponding 95% confidence intervals in **Table 3**. The same weighting and corrections apply as for **Table 2**.

**Table 3.**
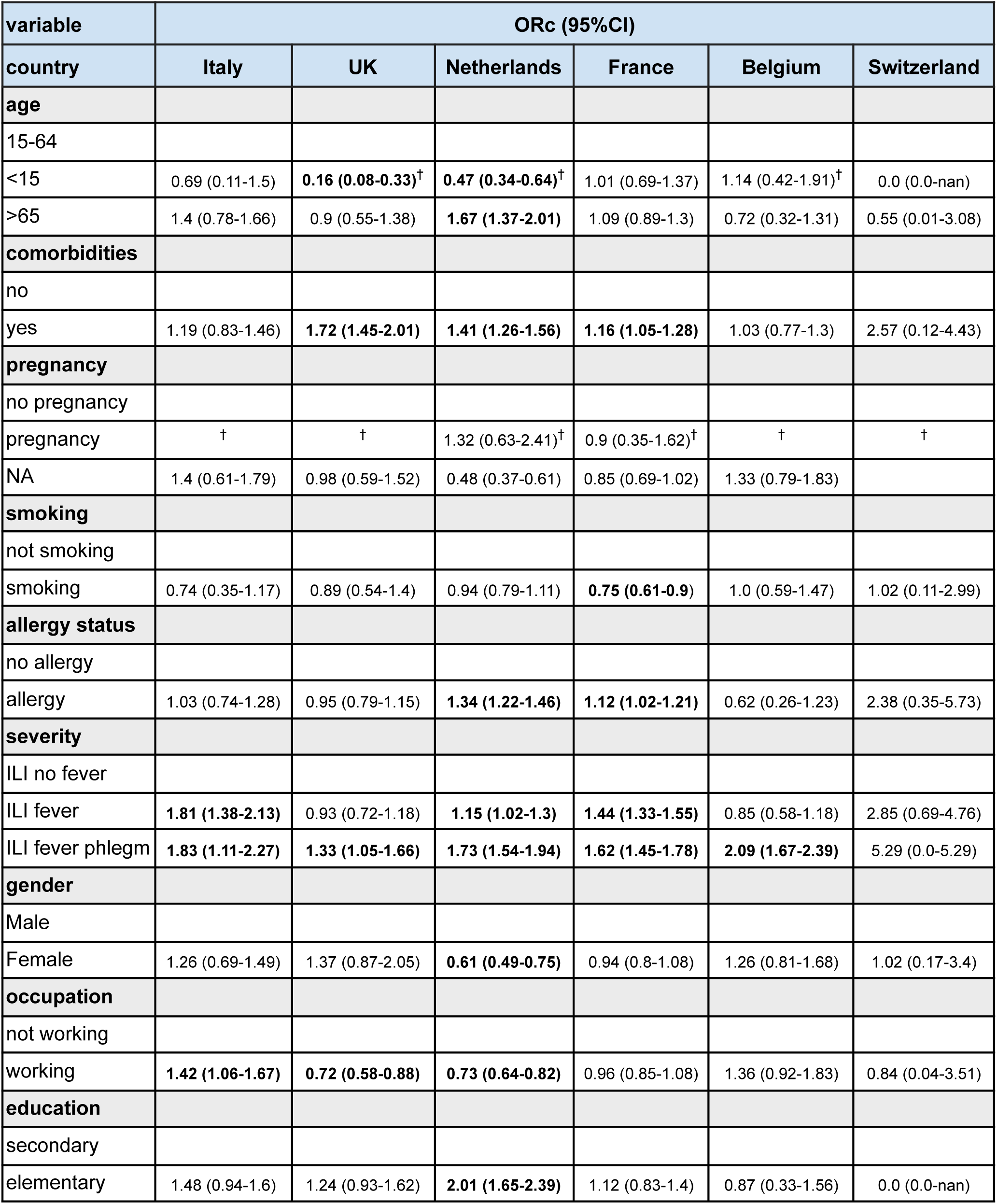
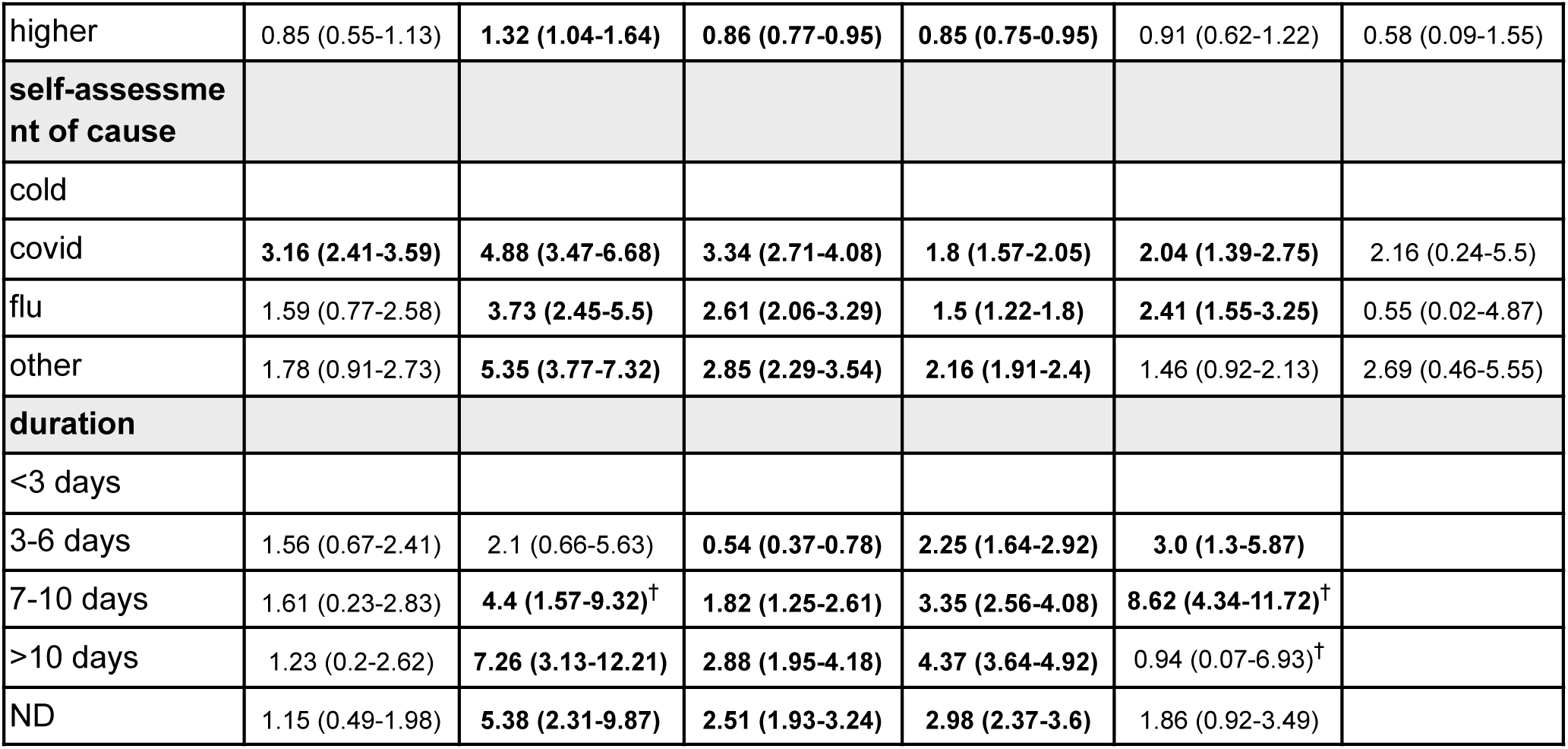
Pandemic period determinants of HCSB. Table shows for each country and category the adjusted odds-ratios and the 95% confidence intervals in parentheses. Reference groups for each covariate are reported at the first row. Only four countries under study collected data during the pre-pandemic period, namely Italy, the UK, France and Switzerland. Odds-ratios are adjusted by the Zhang-Yu correction. Odds-ratios are not computed for groups with too scarce samples. ^†^Group sample lower than 3%. Empty slots represent categories with no cases. ND = No Data. Bold values refer to significant results.

| variable | ORc (95%CI) |  |  |  |  |  |
| --- | --- | --- | --- | --- | --- | --- |
| country | Italy | UK | Netherlands | France | Belgium | Switzerland |
| **age** |  |  |  |  |  |  |
| 15-64 |  |  |  |  |  |  |
| <15 | 0.69 (0.11-1.5) | **0.16 (0.08-0.33)<sup>†</sup>** | **0.47 (0.34-0.64)<sup>†</sup>** | 1.01 (0.69-1.37) | 1.14 (0.42-1.91)<sup>†</sup> | 0.0 (0.0-nan) |
| >65 | 1.4 (0.78-1.66) | 0.9 (0.55-1.38) | **1.67 (1.37-2.01)** | 1.09 (0.89-1.3) | 0.72 (0.32-1.31) | 0.55 (0.01-3.08) |
| **comorbidities** |  |  |  |  |  |  |
| no |  |  |  |  |  |  |
| yes | 1.19 (0.83-1.46) | **1.72 (1.45-2.01)** | **1.41 (1.26-1.56)** | **1.16 (1.05-1.28)** | 1.03 (0.77-1.3) | 2.57 (0.12-4.43) |
| **pregnancy** |  |  |  |  |  |  |
| no pregnancy |  |  |  |  |  |  |
| pregnancy | † | † | 1.32 (0.63-2.41)<sup>†</sup> | 0.9 (0.35-1.62)<sup>†</sup> | † | † |
| NA | 1.4 (0.61-1.79) | 0.98 (0.59-1.52) | 0.48 (0.37-0.61) | 0.85 (0.69-1.02) | 1.33 (0.79-1.83) |  |
| **smoking** |  |  |  |  |  |  |
| not smoking |  |  |  |  |  |  |
| smoking | 0.74 (0.35-1.17) | 0.89 (0.54-1.4) | 0.94 (0.79-1.11) | **0.75 (0.61-0.9)** | 1.0 (0.59-1.47) | 1.02 (0.11-2.99) |
| **allergy status** |  |  |  |  |  |  |
| no allergy |  |  |  |  |  |  |
| allergy | 1.03 (0.74-1.28) | 0.95 (0.79-1.15) | **1.34 (1.22-1.46)** | **1.12 (1.02-1.21)** | 0.62 (0.26-1.23) | 2.38 (0.35-5.73) |
| **severity** |  |  |  |  |  |  |
| ILI no fever |  |  |  |  |  |  |
| ILI fever | **1.81 (1.38-2.13)** | 0.93 (0.72-1.18) | **1.15 (1.02-1.3)** | **1.44 (1.33-1.55)** | 0.85 (0.58-1.18) | 2.85 (0.69-4.76) |
| ILI fever phlegm | **1.83 (1.11-2.27)** | **1.33 (1.05-1.66)** | **1.73 (1.54-1.94)** | **1.62 (1.45-1.78)** | **2.09 (1.67-2.39)** | 5.29 (0.0-5.29) |
| **gender** |  |  |  |  |  |  |
| Male |  |  |  |  |  |  |
| Female | 1.26 (0.69-1.49) | 1.37 (0.87-2.05) | **0.61 (0.49-0.75)** | 0.94 (0.8-1.08) | 1.26 (0.81-1.68) | 1.02 (0.17-3.4) |
| **occupation** |  |  |  |  |  |  |
| not working |  |  |  |  |  |  |
| working | **1.42 (1.06-1.67)** | **0.72 (0.58-0.88)** | **0.73 (0.64-0.82)** | 0.96 (0.85-1.08) | 1.36 (0.92-1.83) | 0.84 (0.04-3.51) |
| **education** |  |  |  |  |  |  |
| secondary |  |  |  |  |  |  |
| elementary | 1.48 (0.94-1.6) | 1.24 (0.93-1.62) | **2.01 (1.65-2.39)** | 1.12 (0.83-1.4) | 0.87 (0.33-1.56) | 0.0 (0.0-nan) |
| higher | 0.85 (0.55-1.13) | **1.32 (1.04-1.64)** | **0.86 (0.77-0.95)** | **0.85 (0.75-0.95)** | 0.91 (0.62-1.22) | 0.58 (0.09-1.55) |
| **self-assessment of cause** |  |  |  |  |  |  |
| cold |  |  |  |  |  |  |
| covid | **3.16 (2.41-3.59)** | **4.88 (3.47-6.68)** | **3.34 (2.71-4.08)** | **1.8 (1.57-2.05)** | **2.04 (1.39-2.75)** | 2.16 (0.24-5.5) |
| flu | 1.59 (0.77-2.58) | **3.73 (2.45-5.5)** | **2.61 (2.06-3.29)** | **1.5 (1.22-1.8)** | **2.41 (1.55-3.25)** | 0.55 (0.02-4.87) |
| other | 1.78 (0.91-2.73) | **5.35 (3.77-7.32)** | **2.85 (2.29-3.54)** | **2.16 (1.91-2.4)** | 1.46 (0.92-2.13) | 2.69 (0.46-5.55) |
| **duration** |  |  |  |  |  |  |
| <3 days |  |  |  |  |  |  |
| 3-6 days | 1.56 (0.67-2.41) | 2.1 (0.66-5.63) | **0.54 (0.37-0.78)** | **2.25 (1.64-2.92)** | **3.0 (1.3-5.87)** |  |
| 7-10 days | 1.61 (0.23-2.83) | **4.4 (1.57-9.32)<sup>†</sup>** | **1.82 (1.25-2.61)** | **3.35 (2.56-4.08)** | **8.62 (4.34-11.72)<sup>†</sup>** |  |
| >10 days | 1.23 (0.2-2.62) | **7.26 (3.13-12.21)** | **2.88 (1.95-4.18)** | **4.37 (3.64-4.92)** | 0.94 (0.07-6.93)<sup>†</sup> |  |
| ND | 1.15 (0.49-1.98) | **5.38 (2.31-9.87)** | **2.51 (1.93-3.24)** | **2.98 (2.37-3.6)** | 1.86 (0.92-3.49) |  |

We recovered the same determinants of HCSB as comorbidities, self-assessed cause of symptoms (here including Covid), severity and duration of symptoms also in the pandemic period consistently across countries. Only Italy differed in the significance of factors of comorbidities and cause of symptoms due to lower population sample in the pandemic with respect to the pre-pandemic period. As for the analysis in the pre-pandemic period, Switzerland did not show significant associations during the pandemic and no conclusions can be derived from this analysis. We found no association of HCSB with female gender individuals in either period. No conclusive results can be derived from the analysis on pregnancy factors.

Our analysis on Italy, UK, France and Switzerland, where pre-pandemic data were available, highlighted that HCSB age determinants changed during the pandemic period in a country-specific fashion and we cannot derive any general trend across countries, whereas working status and education factors remained mostly unaltered in Italy and France. In Italy and France age factors association with HCSB lost significance in the pandemic period, in the UK in the pandemic period ILI patients aged less than 15 years were less likely to seek medical service with respect to the 15-64 age group, whereas no association was estimated in the pre-pandemic period. Age factors in Switzerland remained unaltered and remained non-significant. In general, we found that educational level was not significant in determining HCSB in the pre-pandemic period except for France, while in the pandemic period we found positive association in the UK for the higher education level, and for secondary and higher education levels in the Netherlands. No further significance in the rest of the countries was found for education.

### Influenza-like-illness burden estimations

We use the seasonal ILI burden of cases reported by national health authorities, as collected by national sentinel networks and reported by the FluID and FluNews programmes^23,24^ for comparison with our estimates. In **Figure 2** we show the comparison of the seasonal ILI burden estimated by our model, the *InfluenzaNet* ILI burden (as solid bars) in the three countries that have an ILI case definition that most closely matches the ECDC definition, namely Italy, France and Switzerland (see **Table S1** for the case definitions adopted by these countries).

**Figure 2.**
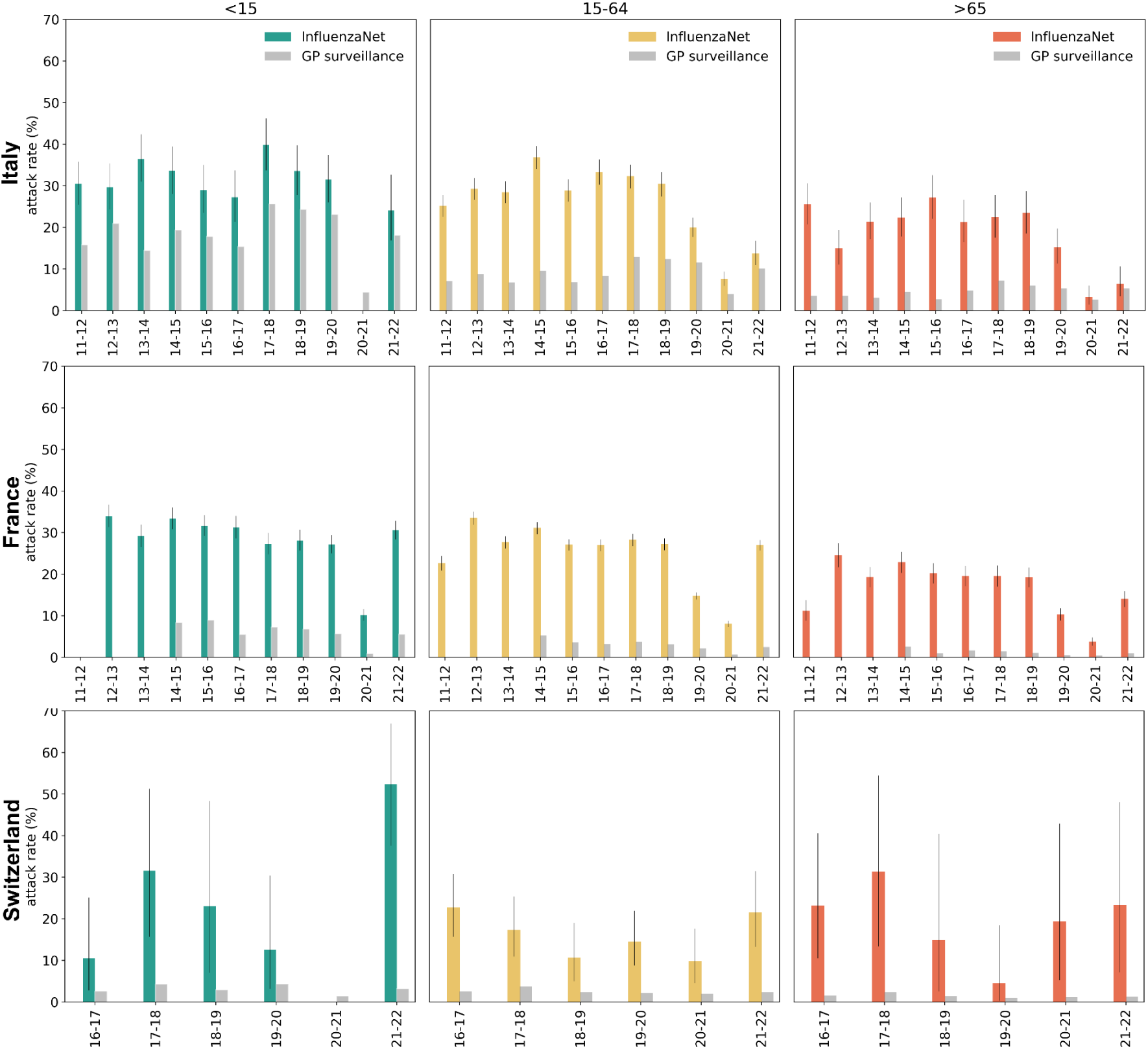
ILI burden for Italy, France and Switzerland. National comparison of InfluenzaNet reported ILI burden, *InfluenzaNet* as colored bars, and *GP surveillance* reported burden as gray bars, across years. Attack rates for ILI cases are expressed in percentage. Whiskers representing the 95%CIs computed through the Clopper-Pearson method. GP surveillance burden collected from ERVISS^25^.

For all countries we found substantially higher burden for all age classes across all seasons with respect to the burden reported by GP surveillance. Still, in Italy our estimations suggest a lower burden of ILI for individuals under 15 years old with respect to previous estimates by *Trentini et al*. (ranging from 2011 to 2020, Supplementary Figure S4), whereas we found higher ILI burden for the adults and elderly age classes, with narrower differences between the two estimates for later seasons. Only in season 2020-2021 we estimate lower burdens for young individuals with respect to GP surveillance.

In France and Switzerland our method systematically estimated a higher ILI burden for all age classes with respect to the reported burden across all seasons. No data for ILI cases were found for Switzerland in the 2020-2021 season for individuals under 15 years old.

In **Figure 3** we show the same comparison in three countries for which the national surveillance system adopts an ILI case definition sensibly different from the ECDC one. Belgium relies on the WHO definition with the addition of sudden onset of symptoms, Netherlands adopts Pel’s criteria^26^, requiring specific temperature measurement, while the UK (England) uses the Royal College of GPs’ Research and Surveillance Centre’s definition (see **Table S1** for the case definitions adopted).

**Figure 3.**
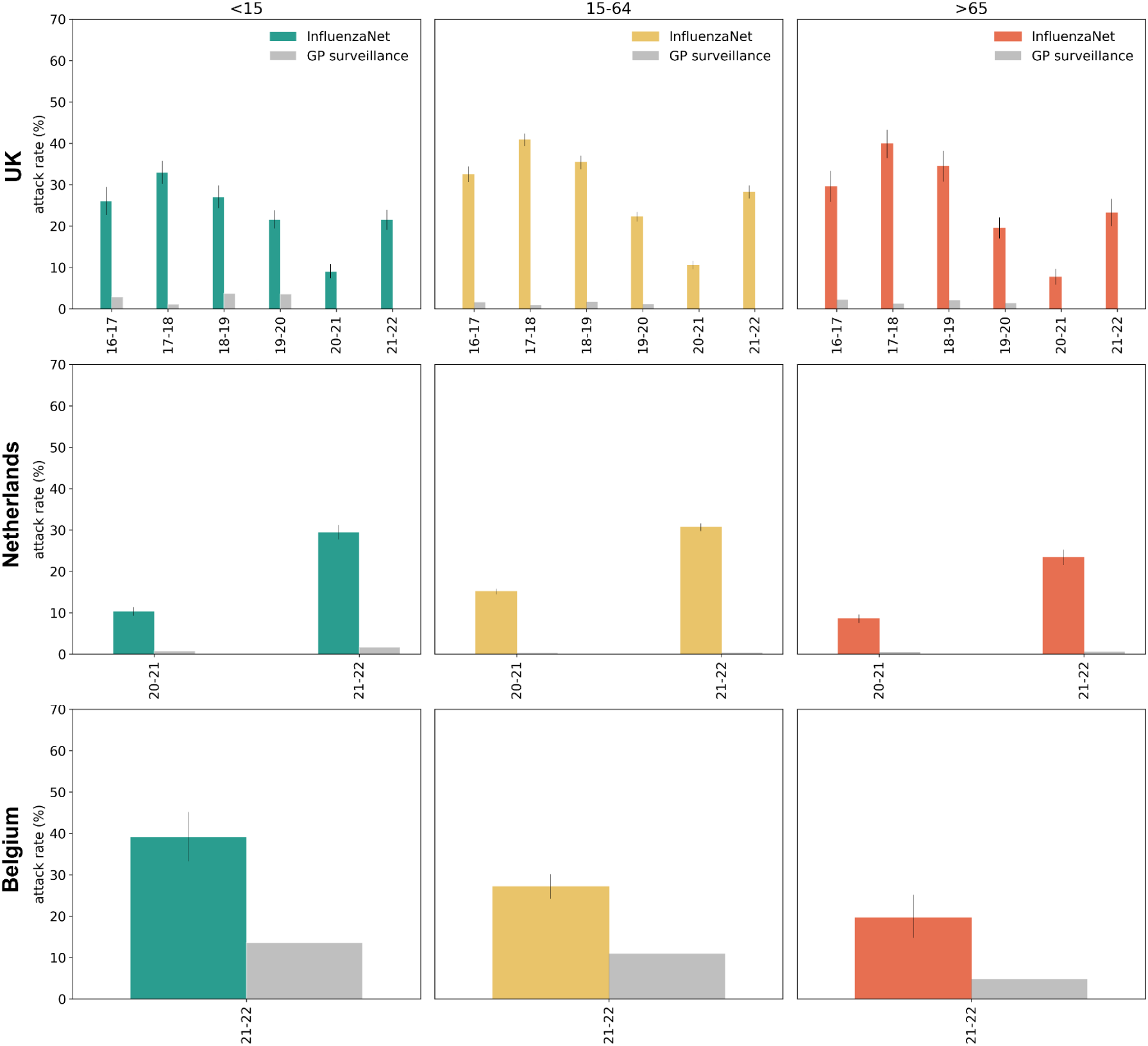
ILI burden for the UK, the Netherlands and Belgium. National comparison of InfluenzaNet reported ILI burden, *InfluenzaNet* as colored bars, and *GP surveillance* reported burden as gray bars, across years. Attack rates for ILI cases are expressed in percentage. Whiskers representing the 95%CIs computed through the Clopper-Pearson method. GP surveillance burden collected from ERVISS^25^.

We found substantially different estimates of ILI burden among the InfluenzaNet participants with respect to the burden reported by national surveillance systems in the UK and the Netherlands, while estimates for Belgium are more consistent with officially reported values. This may be due to the similarity of ILI case definitions of WHO and ECDC, as shown in Figures S2, S3 in a quantitative comparison of the two case definitions for the Italian and French cohorts.

## Discussion

We estimated factors associated with healthcare seeking information for individuals experiencing ILI symptoms in six European countries in pre-pandemic and pandemic seasons, assessing the impact that the pandemic had on specific demographic groups’ medical attendance in all countries. The study was based on information provided by volunteers from the participatory surveillance platform InfluenzaNet. We recovered determinants of healthcare seeking behaviors that are well known in the literature, such as the presence of comorbidities^15^, the duration of symptoms and severity^7^ and self-assessed cause of symptoms (self-diagnosis), previously observed for cold and flu^7^. This aligns with previous approaches, such as the Health Belief Model^27^, linking patients’ health related behavior to their self-diagnosis and perception of their disease susceptibility and its severity^28^, hence confirming the role of psychological factors, though health service access and capacity also warrants consideration. Overall, these results indicate that traditional surveillance systems mainly capture ILI patients reporting severe and/or long lasting symptoms or either believing they had caught a virus, neglecting a wide fraction of population burden who believe they simply caught a cold, or present mild and short-lived symptoms, that still concur to transmission and may result in social and economic activities disruption.

Despite the impact of the pandemic on national healthcare systems and despite national differences between countries in terms of culture, employment rate and national healthcare systems organization, determinants of HCSB that were significant in the pre-pandemic period appeared robust across countries and were not impacted by the pandemic. In specific cases, limited samples of online cohorts, e.g. for Switzerland in both pre-pandemic and pandemic periods and Italy in the pandemic, affected the significance for these factors. Our model suggests that further variables, like age, gender, working status and education level, had a country-dependent impact on HCSB and no general trend can be extracted from results. This heterogeneity was probably influenced by the different burden of Covid-19 infections in demographic groups between countries and by country-specific pandemic policies in terms of healthcare access for working individuals and remote working during the pandemic and the presence of country-dependent medical service in schools and universities^29^. Different age associations with HCSB may be attributed to different Covid-19 burden, testing strategies targeting specific age classes (like required school testing^30–32^) and different pediatric follow up schemes in the pre-pandemic period^33,34^.

By combining reported symptoms and healthcare seeking information, we were able to estimate the burden of ILI for all countries under study. With respect to traditional surveillance systems, this estimate has the advantage to account for ILI patients that typically do not consult healthcare services. Differences between our estimates and the reported burden of ILI across seasons vary from country to country: we observe the lowest underreporting of ILI cases for Italy, France, Switzerland and Belgium, and the highest for the Netherlands and UK, which substantially differ from the reported burden. Beyond different case definitions^35^, this disagreement may reflect the diverse focus of national influenza surveillance systems^36^, more promoted to virological surveillance rather than syndromic surveillance. Moreover, the Netherlands case study only focuses on the pandemic seasons, which are known to have had an impact on the healthcare seeking behavior of ILI patients and on epidemiological data collection^37^. Odds of seeking healthcare shown in **Table 1** for patients self-diagnosing with flu, Covid and other diseases with respect to cold are sensibly higher for the UK cohort than for Italy and France. This possibly points to a lower rate of health care seeking for colds in the UK (and the Netherlands) with respect to Italy and France, which would partly explain the difference in ILI attack rate reported by GP networks in the four countries. Additionally, at the national level, differences in medical service accessibility and healthcare seeking behavior across demographic groups shape surveillance capacity to capture ILI circulation^3^. The pandemic stressed national health systems and, despite the attempts to implement remote medical services, stay-at-home orders, social distancing recommendations and sensibilization to prevention and control of the disease may have impacted access to health-care centers for specific demographic groups in all countries.

Previous studies adopted multiplier approaches to correct reported burden of ILI for healthcare seeking behavior^10,38,39^. These studies focused on single countries, hence relying on country-specific ILI case definitions. When studying multiple countries using different case definitions, a multiplier approach based on a single case definition would still lead to inconsistent comparisons. In this work we adopted the ECDC definition of influenza-like-illness^1^ in order to study healthcare seeking behavior in a consistent way in all the six countries and get a cross-country picture of ILI dynamics across seasons. Historically, national surveillance systems relied on their own specific case definitions^35^, and even if there have been efforts to standardize ILI definitions at the international level^40^, heterogeneities persist (**Table S1**).

The use of digital cohorts allowed us to control for a wide variety of socio-demographic information at a high temporal resolution and employ a consistent ILI case definition across all countries under study and across seasons. This information is typically not collected by traditional surveillance systems and can suggest important insights in how ILI patients refer to healthcare service. Most importantly, this information can uncover the burden of disease affecting a wide fraction of the population experiencing milder and short-lived symptoms of influenza. Additionally, the harmonized data collection supported by the digital platforms can mitigate historical country level heterogeneities of surveillance systems, like ILI case definitions, providing consistent burden comparisons and possible interventions at European level.

As a further point, these tools are essential to study the temporal evolution of population behavior and their drivers, which may change drastically during pandemics^41^, beyond national borders. For these reasons, participatory surveillance systems, like InfluenzaNet, FluNearYou (United States)^18^ or FluTracking (New Zealand)^19^, constitute critical tools that enrich national surveillance systems, especially in countries where ILI cases surveillance is neglected.

## Limitations

Healthcare seeking behavior varied substantially across countries, both when looking at the overall period and in the pre-pandemic and pandemic periods separately. However, the cause for this heterogeneity at national level must be searched with caution, since on one side this could reflect national specific health-care system organization, but it could also hide the different cultural or health-related habits of the population engaged in the online participatory platform across countries.

Biases may be present both on estimates from the online participatory surveillance platform and on reported burden from sentinel surveillance. On one hand, reported ILI burden are computed on sentinel networks consultation rates, representing the share of patients with ILI among patients contacting the sentinel network on a weekly basis, burden may be under-estimated due to healthcare seeking behavior of ILI patients, i.e. patients suffering from ILI but not contacting medical service, but also over-estimated in specific weeks if HCSB from ILI patients grows substantially with respect to HCSB of general consulting patients; on the other hand, while estimates of ILI burden from the online cohorts do not depend on HCSB, online cohorts may not be representative of the population behavior since these may represent individuals that are more attentive to symptoms, tend to report their health status and to seek medical service more often than the average patient.

## Conclusions

Our work highlights that despite differences in culture, socio-economic and demographic structure, healthcare systems and participation in the study, comorbidities, self-assessed cause of symptoms, duration and severity of symptoms drove healthcare seeking behavior consistently across countries.

Our approach, leveraging online digital cohorts, shows how participatory surveillance systems can be exploited to complement traditional surveillance in estimating important public health aspects, in this specific case influenza burden among the general population, in a consistent fashion across countries^21,42–44^.

By directly probing the general population about their healthcare seeking behavior, these tools can help overcome an important limitation of traditional surveillance, namely the reliance on individuals seeking healthcare assistance to assess the epidemiological situation among the general population.

The adoption of online participatory surveillance systems in further EU and non-EU countries may help complement the activities of national surveillance systems.

## Methods

### InfluenzaNet data

We rely on InfluenzaNet^20^, a multi-country and harmonized online participatory surveillance system collecting data on symptoms of diseases like Influenza and Covid-19. InfluenzaNet is based on a voluntary weekly survey focused on tracking symptoms of respiratory diseases and participants’ behavior and health habits. It provides estimates of ILI incidence, insights on vaccination and testing attitudes, and measures of behavioral response to health threats. The project was born in the year 2003 and is currently available in 11 countries.

InfluenzaNet^20^ is supported by several European projects through a multi-country academic initiative (EPIPOSE^45^, PANDEM-2, VERDI^46^). We analyze the data collected from the surveys on self-reported ILI symptoms and patients’ behavior towards healthcare seeking. We post-stratify the InfluenzaNet population to correct for the respective national populations structure by age and sex and control for the survey sampling biases. We employ a logistic regression to explain healthcare seeking behavior in terms of demographics, medical conditions and symptoms characteristics, accounting for control variables like educational level, personal belief of symptoms cause and working status.

Individuals can register on their InfluenzaNet country-specific websites and fill in the intake questionnaire regarding their personal information, e.g. year of birth, gender, education and medical conditions among others. Once completed, users can fill in the weekly survey regarding their symptoms. The survey ends if no symptoms are reported in the week, otherwise the survey develops into further questions on the onset of symptoms, medical care, adoption of social distancing measures among others^17^.

National surveys may develop customized versions removing or including specific questions, but all share the same questions indexes and texts, which is crucial to harmonize data collection and analysis. National platforms were developed at different times, hence data coverage in time across countries may vary: in Italy, UK and France data collection started in 2011, in the Netherlands in 2020, in Belgium in 2021, in Switzerland in 2016. Since participation occurs on a voluntary basis, online cohorts are not representative of the underlying national population and post-stratification methods are necessary.

### Data filtering and treatment

We removed repeated weekly and intake surveys, i.e. surveys filled more than once by the same user in the same week, keeping the last report of the week in order to have the most up-to-date submissions. We considered users who filled in at least two symptoms surveys in the whole time period of analysis in order to minimize noise due to occasional users’ submissions, consistently with previous studies^47,48^.

We discarded weekly survey submissions not reporting symptoms compatible with the ECDC definition of ILI^1^. In order to correctly assess if the ILI user sought medical service in the following weeks, we grouped ILI episodes that were prolonged in time and merged them into a single episode with all the necessary information on symptoms, onset week, healthcare seeking behavior and user’s metadata, i.e. age, gender, comorbidities, smoking status, allergies, pregnancy, working status and education level.

We merged the weekly data with the intake data by associating to each submission user their most recent intake survey, in order to always consider their most up-to-date medical condition. In some countries, e.g. the UK, the intake survey must be filled every year by the user before being able to submit new symptoms answers, in other cases, e.g. in Italy, the web page of the weekly survey shows a dedicated reminder every year (move to limitations).

### Target and control variables Age class

We aggregated participatory surveillance data in three age classes, i.e. *<15*, *15-64*, *>65* years old to allow comparison with those used by the ECDC and WHO Global Influenza Programme in the reported ILI burden in the population^23,24^.

### Gender

Participants were asked to report their gender among “Male”, “Female” and “Other”, however the third option was only recently adopted in some countries and, where available, only 1% of participants identified as “Other”. For sake of sampling and consistency across countries we focused only on the “Male” and “Female” options.

### Educational level

We aggregated educational levels across countries to *elementary*, *secondary* and *higher* levels. We assigned users who declared still being a student but reported no educational title, users who only declared not having any degree and users who declared having completed an elementary grade to the *elementary* level. We assigned users who have completed intermediate school or high school to *secondary*, those who have a bachelor, master or PhD title to *higher* grade.

### Working status

To discriminate between those who are actively working (and may benefit of job-related healthcare accessibility) and those who are not, we aggregated occupational status to two statuses, respectively assigning unemployed, students, homemakers, on leave, retired or other to *not working* and those who report being full-time, part-time and self-employed to *working*.

### Medical condition: comorbidities, smoking status, allergy and pregnancy

We labeled users regularly taking medications for immunosuppression, asthma, other pulmonary chronic conditions, diabetes, cardiovascular or kidney diseases as *yes*, and those who did not report any of the above as *no*.

We consider individuals who suffer from allergies causing respiratory symptoms like hay fever, dust mite, pets hair or other as *allergy*, those reported smoking no matter how frequently as *smoking*, whereas absence of each of the above conditions results in classification to *no allergy*, *no smoking*. Women in the 15-64 age class who reported being pregnant were assigned to *pregnancy*, while those not reporting pregnancy were assigned to *no pregnancy*, women in the other two age classes and males were assigned to *NA (not applying)*.

### Duration of symptoms

We measured duration of symptoms by taking the declared date of end of symptoms and onset of symptoms (fever onset if missing) and aggregate into four ranges, *<3 days*, *3-6 days*, *7-10 days*, *>10 days* and *ND* (ND= No Data) when the information was not provided.

### Severity of symptoms

Following the ECDC ILI definition^1^, we distinguish three levels of symptom severity, *ILI no fever*, *ILI fever* and *ILI fever phlegm*, respectively describing ILI symptoms without fever, with fever and those with fever and shortness of breath, consistently with previous studies^7^. ARI definition requires sudden onset and one respiratory symptom, hence all ILI episodes are also ARI by definition. To avoid ill-defined regression, we classify episodes belonging to the highest severity class only, e.g. patient reporting sudden onset, cough and fever is classified as *ILI fever* and not as *ARI*.

### Self-assessment of cause

We considered the patient’s self-diagnosis, hence the patient’s belief of the symptoms’ cause, accounting for *cold*, *flu*, *Covid-19* or *other*.

### Healthcare seeking

Finally, we considered whether the user reporting the ILI episode had ever sought medical service, either in person or remotely, in the weeks following the onset of symptoms as *medical service* or *no medical service*. Medical service includes consulting GPs, GP’s nurses, hospital admission, emergency departments and other medical services. Remote consultation can occur via telephone or Internet. This played as our dependent variable for the regression.

### Survey population weighting

In order to adjust the statistical representativeness of the InfluenzaNet participants by age class and sex, we weighed respondents’ answers accounting for their sampling with respect to their expected population structure as reported by national statistical offices.

We collected the official statistics to obtain the national population structure by age class and sex (ISTAT for Italy, ONS for the UK,,CBS for the Netherlands, the French National Institute of Statistics and Economic Studies (INSEE) for France, StatBel for Belgium, Federal Statistical Office for Switzerland). For each country, we computed the weights for our population of age *a*, gender *g* in season *s* as:

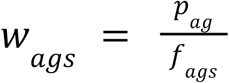

where *p* is the reference population fraction in age class *a* and sex *g* reported by the national statistical institute of the respective country in the reference year, and *f* is the fraction of InfluenzaNet participants in age class *a* and gender *g* in season *s*. We used the resulting weights *w_ags_* to weigh the survey responses to perform the logistic regression and compute the InfluenzaNet endogenous burden.

### Zhang-Yu correction

Odds-ratios represent the odds of observing a given outcome in an exposed group with respect to the odds of observing the same outcome within a reference group. They are used in case-control studies, where the risk-ratio cannot be computed since the population at the denominator is unknown or non-representative of the expected population. Odds-ratios are computed by logistic regressions in case-control studies and are considered approximations of the risk-ratio (computed in cohort studies by log-binomial regression) when the incidence of the outcome variable is rare (<10%). This is never the case in any of the countries under study, as we see in **Table 1**. To adjust the odds-ratios computed by the logistic regression for all covariates, we use the Zhang-Yu correction, formalized as:

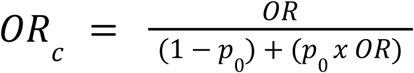

which considers the incidence *p*_0_ of healthcare seeking behavior in the reference group^49,50^, while *OR* is the odds-ratio computed by the logistic regression for any covariate.

### Reported ILI burden national estimates from GP surveillance

We obtained the reported ILI burden for each country from the FluID and FluNews websites^23,24^. These platforms report the consultation rates reported by national public health agencies gathered by the respective sentinel network of general practitioners and pediatrics. The data comes as aggregated in four age classes, namely *0-4*, *5-14*, *15-64*, *>65* years old. Among the InfluenzaNet population the *0-4* years old sample is too scarce, hence for sake of comparison we aggregated the reported statistics in the three age classes, i.e. *<15*, *15-64*, *>65* years old.

### InfluenzaNet reported ILI burden

The model estimating ILI burden computes the fraction of ILI episodes within our digital cohorts. Since participants change across weeks, the denominator of our ILI incidence is not fixed in time. We computed the fraction of ILI episodes occurring in each season among season participants. For each participant in each season we account for a single ILI episode, if any, while multiple ILI episodes are discarded. Seasonal incidences are adjusted by age and sex (see Section *Survey population weighting*) to obtain the seasonal ILI burden. We defined it as *B*^*I*^ _*as*_.

### Ethics Declaration

Grippenet/Covidnet was reviewed and approved by the French Advisory Committee for research on information treatment in the health sector (CCTIRS), and by the French National Commission on Informatics and Liberty (CNIL) - the authorities ruling on all matters related to ethics, data, and privacy in France.

The research protocol of Infectieradar was shared with the Medical Ethics Review Committee Utrecht, and an official waiver for ethical approval (reference number: WAG/avd/20/008757; protocol 20-131) was obtained given the nature of data collection. Participants of Infectieradar had to agree to the privacy statement upon registration, which described the processing of personal data and research results, website security measures taken, and how to file a complaint. Additionally, they had to give consent to participate in the study. Participants were eligible to withdraw from the study at any time. Persons had to be 16 years or over to be able to participate.

For influweb.it the research was conducted in agreement with Italian regulations on privacy and data collection and treatment. The institutional review board of ISI Foundation, upon consultation with the Italian Data Protection authority, waived the ethical approval for this study.

Infectierdar Belgium has obtained ethical approval from the UZA Ethics Committee and the University of Hasselt Medical Ethics Committee of Belgium (Belgian registration number: B300202000227 with reference 20/41/539). Infectieradar.be processes personal data in a lawful manner, in accordance with the law and the GDPR regulations. You can read more about this in the Informed Consent Form.

Grippenet data is handled in accordance with Swiss law on data protection and privacy. UK FluSurvey is a national surveillance scheme. Participants accept a privacy statement on registration, describing data processing and consenting to the use of their data for these purposes. Participants are eligible to withdraw from the study at any time.

## Data Availability

Individual data collected for this study cannot be publicly shared due to privacy requirements.

## Data Sharing

Individual data collected for this study cannot be publicly shared due to privacy requirements.

## Acknowledgements

M.M., L.H., N. H., D.P. and V.C. acknowledge funding from the EU Commission in the framework of the EU Horizon Project VERDI. VC acknowledges funding from the EU Horizon 2020 grants MOOD (H2020-874850).

M.M., L.H., N.H., V.C. and D.P. acknowledge support by the ESCAPE project (101095619), funded by the European Union. Views and opinions expressed are however those of the author(s) only and do not necessarily reflect those of the European Union or European Health and Digital Executive Agency (HADEA). Neither the European Union nor the granting authority can be held responsible for them.

M.M., L.H., N.H. and D.P. have received funding from the European Union’s Horizon 2020 research and innovation programme - project EpiPose (Grant agreement number 101003688). This work reflects only the authors’ view. The European Commission is not responsible for any use that may be made of the information it contains.

Infectieradar is supported by the Ministry of Health, Welfare and Sports (VWS), of the Netherlands.

Infectieradar.be is supported by the Belgian Scientific Institute of Public Health, Sciensano. Grippenet.ch is funded by the Institute of Global Health, University of Geneva, Switzerland. Grippenet/Covidnet is partially supported by Santé publique France.

FluSurvey is funded by the UK Health Security Agency and was previously funded by Public Health England.

## Authors contributions

M.M. and D.P. have directly accessed and verified the underlying data reported in the manuscript. All authors had full access to all the data in the study and accept responsibility to submit for publication. All authors participated in the data collection. M.M. conducted the statistical analysis. M.M. and D.P. wrote the manuscript. M.M., N.G. and D.P. designed the research. All authors participated in critically reviewing the analysis and the final version of the manuscript and interpreting the findings.

## References

1 EU case definitions. 2018; published online June 7. https://www.ecdc.europa.eu/en/all-topics/eu-case-definitions (accessed Feb 12, 2024).

2 Giacchetta I, Primieri C, Cavalieri R, Domnich A, de Waure C. The burden of seasonal influenza in Italy: A systematic review of influenza-related complications, hospitalizations, and mortality. Influenza Other Respir Viruses 2022; 16: 351–65.

3 Moss R, Zarebski AE, Carlson SJ, McCaw JM. Accounting for Healthcare-Seeking Behaviours and Testing Practices in Real-Time Influenza Forecasts. Trop Med Infect Dis 2019; 4: 12.

4 Iuliano AD, Roguski KM, Chang HH, et al. Estimates of global seasonal influenza-associated respiratory mortality: a modelling study. The Lancet 2018; 391: 1285–300.

5 Van Cauteren D, Vaux S, de Valk H, Le Strat Y, Vaillant V, Lévy-Bruhl D. Burden of influenza, healthcare seeking behaviour and hygiene measures during the A(H1N1)2009 pandemic in France: a population based study. BMC Public Health 2012; 12: 947.

6 Ariza M, Guerrisi C, Souty C, et al. Healthcare-seeking behaviour in case of influenza-like illness in the French general population and factors associated with a GP consultation: an observational prospective study. BJGP Open 2018; 1. DOI:10.3399/bjgpopen17X101253.

7 Peppa M, John Edmunds W, Funk S. Disease severity determines health-seeking behaviour amongst individuals with influenza-like illness in an internet-based cohort. BMC Infect Dis 2017; 17: 238.

8 Baltrusaitis K, Reed C, Sewalk K, Brownstein JS, Crawley AW, Biggerstaff M. Healthcare-Seeking Behavior for Respiratory Illness Among Flu Near You Participants in the United States During the 2015–2016 Through 2018–2019 Influenza Seasons. J Infect Dis 2020; 226: 270–7.

9 Meng H, Liao Q, Suen LKP, O’Donoghue M, Wong CM, Yang L. Healthcare seeking behavior of patients with influenza like illness: comparison of the summer and winter influenza epidemics. BMC Infect Dis 2016; 16: 499.

10 Wang X, Wu S, Yang P, et al. Using a community based survey of healthcare seeking behavior to estimate the actual magnitude of influenza among adults in Beijing during 2013-2014 season. BMC Infect Dis 2017; 17: 120.

11 Zhang Q, Feng S, Wong IOL, Ip DKM, Cowling BJ, Lau EHY. A population-based study on healthcare-seeking behaviour of persons with symptoms of respiratory and gastrointestinal-related infections in Hong Kong. BMC Public Health 2020; 20: 402.

12 Sun X, Luo S, Lou L, et al. Health seeking behavior and associated factors among individuals with cough in Yiwu, China: a population-based study. BMC Public Health 2021; 21: 1157.

13 Rolfes MA, Foppa IM, Garg S, et al. Annual estimates of the burden of seasonal influenza in the United States: A tool for strengthening influenza surveillance and preparedness. Influenza Other Respir Viruses 2018; 12: 132–7.

14 Frey A, Tilstra AM, Verhagen MD. Inequalities in healthcare use during the COVID-19 pandemic. Nat Commun 2024; 15: 1894.

15 Chawla D, Benitez A, Xu H, et al. Predictors of Seeking Care for Influenza-Like Illness in a Novel Digital Study. Open Forum Infect Dis 2022; 10: ofac675.

16 Mølbak K, Widgren K, Jensen KS, et al. Burden of illness of the 2009 pandemic of influenza A (H1N1) in Denmark. Vaccine 2011; 29: B63–9.

17 Koppeschaar CE, Colizza V, Guerrisi C, et al. Influenzanet: Citizens Among 10 Countries Collaborating to Monitor Influenza in Europe. JMIR Public Health Surveill 2017; 3: e7429.

18 Flu Near You. End. Pandemics. https://endingpandemics.org/projects/flu-near-you/ (accessed Feb 16, 2024).

19 New Zealand Reports | Flutracking.net. https://info.flutracking.net/reports/new-zealand-reports/ (accessed Feb 16, 2024).

20 InfluenzaNet Project. https://influenzanet.info/explore-data (accessed Feb 12, 2024).

21 Guerrisi C, Turbelin C, Blanchon T, et al. Participatory Syndromic Surveillance of Influenza in Europe. J Infect Dis 2016; 214: S386–92.

22 Clopper CJ, Pearson ES. The Use of Confidence or Fiducial Limits Illustrated in the Case of the Binomial. Biometrika 1934; 26: 404–13.

23 Flu News Europe | Primary care data. https://flunewseurope.org/PrimaryCareData/ILI (accessed Feb 12, 2024).

24 Influenza surveillance outputs. https://www.who.int/teams/global-influenza-programme/surveillance-and-monitoring/influenza-surveillance-outputs (accessed Feb 12, 2024).

25 The European Respiratory Virus Surveillance Summary (ERVISS). 2025; published online April 11. https://www.ecdc.europa.eu/en/publications-data/european-respiratory-virus-surveillance-summary-erviss (accessed April 15, 2025).

26 Pel J. Proefonderzoek naar de frequentie en de aetiologie van griepachtige ziekten in de winter 1963-1964. Huisarts en Wetenschap 1965; 8: 4.

27 Abraham SA, Agyare DF, Yeboa NK, et al. The Influence of COVID-19 Pandemic on the Health Seeking Behaviors of Adults Living With Chronic Conditions: A View Through the Health Belief Model. J Prim Care Community Health 2023; 14: 21501319231159459.

28 Oberoi S, Chaudhary N, Patnaik S, Singh A. Understanding health seeking behavior. J Fam Med Prim Care 2016; 5: 463–4.

29 Jansen DE, Visser A, Vervoort JP, et al. School and adolescent health services in 30 European countries: a description of structure and functioning, and of health outcomes and costs. Brussels: European Commission, 2018 https://www.drugsandalcohol.ie/39968/ (accessed April 2, 2024).

30 Mass asymptomatic COVID-19 testing in schools, colleges and HE institutions. GOV.UK. https://www.gov.uk/government/publications/mass-asymptomatic-covid-19-testing-in-schools-colleges-and-he-institutions (accessed April 2, 2024).

31 Coronavirus (COVID-19) asymptomatic testing in schools and colleges. GOV.UK. 2021; published online July 8. https://www.gov.uk/government/publications/coronavirus-covid-19-asymptomatic-testing-in-schools-and-colleges/coronavirus-covid-19-asymptomatic-testing-in-schools-and-colleges (accessed April 2, 2024).

32 COVID-19 | European School of Bergen (NL). https://www.esbergen.eu/covid-19/ (accessed April 2, 2024).

33 Biasci P, Sanz AC, Pop TL, Pettoello-Mantovani M, D’Avino A, Nigri L. The State of Children’s Health in Europe. J Pediatr 2019; 209: 260–261.e1.

34 Ehrich J, Namazova-Baranova L, Pettoello-Mantovani M. Introduction to “Diversity of Child Health Care in Europe: A Study of the European Paediatric Association/Union of National European Paediatric Societies and Associations”. J Pediatr 2016; 177: S1–10.

35 Aguilera JF, Paget WJ, Mosnier A, et al. Heterogeneous case definitions used for the surveillance of influenza in Europe. Eur J Epidemiol 2003; 18: 751–4.

36 de Fougerolles TR, Damm O, Ansaldi F, et al. National influenza surveillance systems in five European countries: a qualitative comparative framework based on WHO guidance. BMC Public Health 2022; 22: 1151.

37 Seasonal influenza - Annual Epidemiological Report for 2020-2021. 2021; published online Aug 26. https://www.ecdc.europa.eu/en/publications-data/seasonal-influenza-annual-epidemiological-report-2020-2021 (accessed April 15, 2025).

38 Reed C, Chaves SS, Kirley PD, et al. Estimating Influenza Disease Burden from Population-Based Surveillance Data in the United States. PLOS ONE 2015; 10: e0118369.

39 Biggerstaff M, Reed C, Epperson S, et al. Estimates of the Number of Human Infections With Influenza A(H3N2) Variant Virus, United States, August 2011–April 2012. Clin Infect Dis 2013; 57: S12–5.

40 Fitzner J, Qasmieh S, Mounts AW, et al. Revision of clinical case definitions: influenza-like illness and severe acute respiratory infection. Bull World Health Organ 2018; 96: 122–8.

41 Eales O, Teo M, Price DJ, et al. Temporal trends in test-seeking behaviour during the COVID-19 pandemic. Math Med Life Sci; 0: 2521858.

42 Paolotti D, Carnahan A, Colizza V, et al. Web-based participatory surveillance of infectious diseases: the Influenzanet participatory surveillance experience. Clin Microbiol Infect 2014; 20: 17–21.

43 Perrotta D, Tizzoni M, Paolotti D. Using Participatory Web-based Surveillance Data to Improve Seasonal Influenza Forecasting in Italy. In: Proceedings of the 26th International Conference on World Wide Web. Perth Australia: International World Wide Web Conferences Steering Committee, 2017: 303–10.

44 Ten-year performance of Influenzanet: ILI time series, risks, vaccine effects, and care-seeking behaviour - ScienceDirect. https://www.sciencedirect.com/science/article/pii/S1755436515000638 (accessed Nov 14, 2023).

45 Epidemic intelligence to minimize 2019-nCoV’s public health, economic and social impact in Europe | EpiPose | Project | Fact sheet | H2020 | CORDIS | European Commission. https://cordis.europa.eu/project/id/101003688 (accessed Feb 12, 2024).

46 Home - VERDI PROJECT. https://verdiproject.org/ (accessed Feb 12, 2024).

47 Guerrisi C, Turbelin C, Souty C, et al. The potential value of crowdsourced surveillance systems in supplementing sentinel influenza networks: the case of France. Eurosurveillance 2018; 23: 1700337.

48 Perrotta D, Bella A, Rizzo C, Paolotti D. Participatory Online Surveillance as a Supplementary Tool to Sentinel Doctors for Influenza-Like Illness Surveillance in Italy. PLOS ONE 2017; 12: e0169801.

49 Debin M, Colizza V, Blanchon T, Hanslik T, Turbelin C, Falchi A. Effectiveness of 2012–2013 influenza vaccine against influenza-like illness in general population. Hum Vaccines Immunother 2014; 10: 536–43.

50 Zhang J, Yu KF. What’s the Relative Risk?A Method of Correcting the Odds Ratio in Cohort Studies of Common Outcomes. JAMA 1998; 280: 1690–1.

