## SupplementaryMaterial for "Healthcare-seeking behavior and hidden influenza-like-illness across the COVID-19 pandemic: a multi-country participatory surveillance study"

**Episode based healthcare seeking behavior**

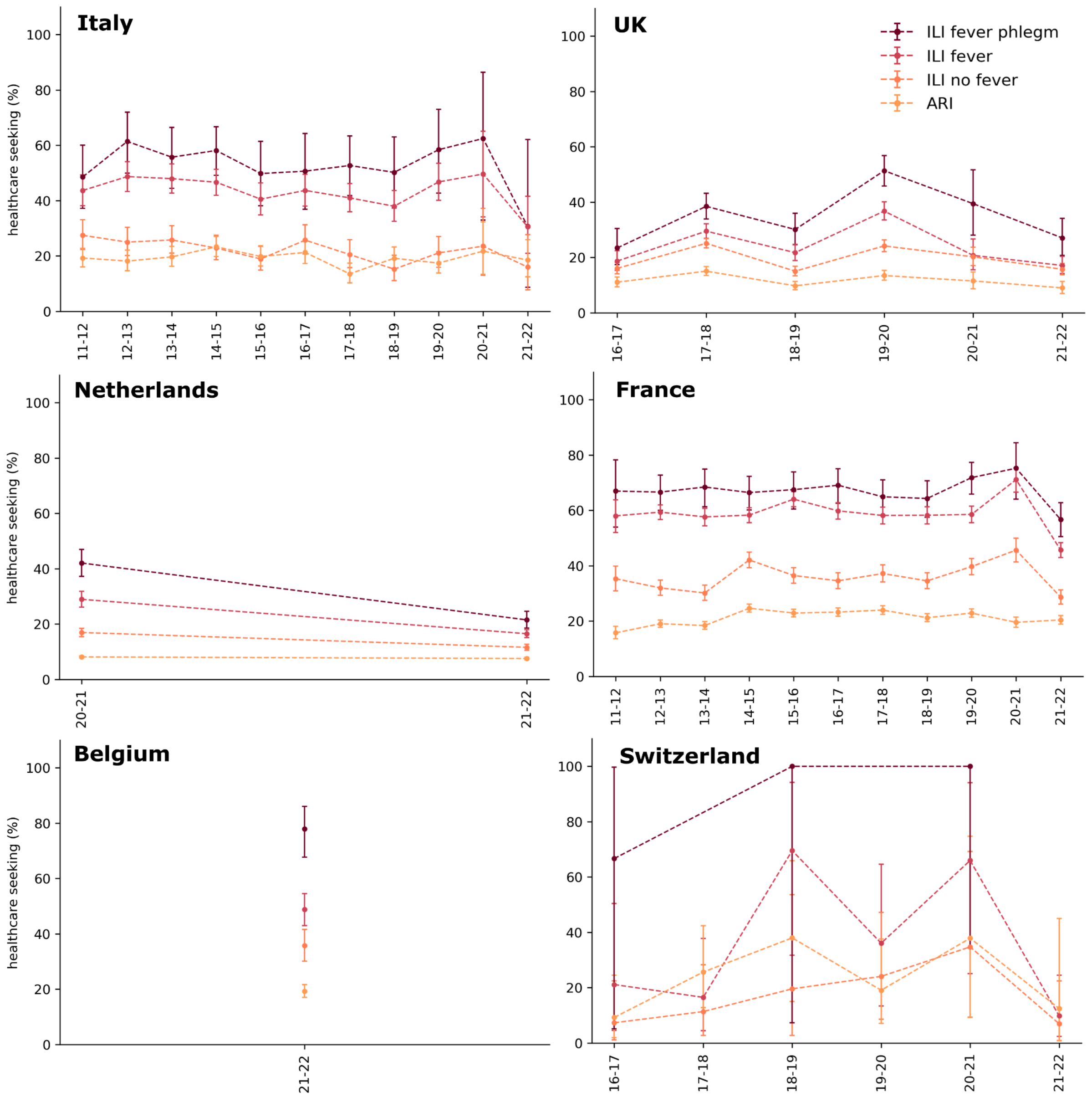


**Supplementary Figure S1. Episode based healthcare seeking behavior by severity of symptoms.** Adjusted healthcare seeking rates of ILI episodes from online cohorts in the six countries under study. Multiple ILI episodes from the same participants are considered independent. Rates are adjusted by age and sex of participants. Darker colors represent more severe symptoms, clearer colors represent less severe symptoms. ARI (Acute Respiratory Infection) follows the ECDC case definition. Whiskers represent the 95% confidence interval computed through the Clopper-Pearson method.

**Unadjusted descriptive statistics**

| variable | % of ILI episodes (occurrence) | | | | | |
|---|---|---|---|---|---|---|
| country | Italy | UK | Netherlands | France | Belgium | Switzerland |

| | | | | | | |
|---|---|---|---|---|---|---|
| **medical service** | | | | | | |
| No | 61.0 (3408) | 78.0 (8527) | 83.0 (7945) | 55.0 (10599) | 60.0 (289) | 78.0 (136) |
| Yes | 39.0 (2170) | 22.0 (2445) | 17.0 (1663) | 45.0 (8588) | 40.0 (191) | 22.0 (38) |
| **age class** | | | | | | |
| <15 | 9.0 (519) | 2.0 (233) | 1.0 (84) | 7.0 (1297) | 3.0 (13) | 8.0 (14) |
| 15-64 | 83.0 (4621) | 74.0 (8113) | 84.0 (8101) | 75.0 (14483) | 70.0 (337) | 79.0 (138) |
| >65 | 8.0 (438) | 24.0 (2626) | 15.0 (1423) | 18.0 (3407) | 27.0 (130) | 13.0 (22) |
| **gender** | | | | | | |
| Female | 49.0 (2717) | 71.0 (7843) | 71.0 (6780) | 68.0 (13079) | 72.0 (344) | 68.0 (119) |
| Male | 51.0 (2861) | 29.0 (3129) | 29.0 (2828) | 32.0 (6108) | 28.0 (136) | 32.0 (55) |
| **occupation** | | | | | | |
| not working | 43.0 (2409) | 39.0 (4329) | 31.0 (3015) | 46.0 (8813) | 41.0 (198) | 32.0 (55) |
| working | 57.0 (3169) | 61.0 (6643) | 69.0 (6593) | 54.0 (10374) | 59.0 (282) | 68.0 (119) |
| **education** | | | | | | |
| elementary | 22.0 (1248) | 13.0 (1404) | 3.0 (318) | 12.0 (2247) | 5.0 (26) | 13.0 (23) |
| higher | 42.0 (2353) | 62.0 (6784) | 58.0 (5576) | 61.0 (11612) | 72.0 (348) | 59.0 (103) |
| secondary | 35.0 (1977) | 25.0 (2784) | 39.0 (3714) | 28.0 (5328) | 22.0 (106) | 28.0 (48) |
| **duration** | | | | | | |
| <3 days | 5.0 (270) | 2.0 (242) | 8.0 (792) | 3.0 (636) | 6.0 (27) | 5.0 (9) |
| 3-6 days | 14.0 (768) | 5.0 (569) | 13.0 (1249) | 9.0 (1818) | 9.0 (42) | 17.0 (29) |
| 7-10 days | 4.0 (225) | 2.0 (267) | 4.0 (411) | 3.0 (650) | 2.0 (10) | 7.0 (13) |
| >10 days | 3.0 (184) | 4.0 (416) | 3.0 (244) | 3.0 (588) | 1.0 (5) | 5.0 (9) |
| ND | 74.0 (4131) | 86.0 (9478) | 72.0 (6912) | 81.0 (15495) | 82.0 (396) | 66.0 (114) |
| **co-morbidities** | | | | | | |
| yes | 20.0 (1104) | 27.0 (2971) | 26.0 (2474) | 25.0 (4746) | 54.0 (260) | 13.0 (22) |
| no | 80.0 (4474) | 73.0 (8001) | 74.0 (7134) | 75.0 (14441) | 46.0 (220) | 87.0 (152) |
| **pregnancy** | | | | | | |
| NA | 59.0 (3305) | 45.0 (4934) | 38.0 (3635) | 46.0 (8748) | 47.0 (227) | 44.0 (76) |
| no pregnancy | 40.0 (2242) | 55.0 (6004) | 62.0 (5926) | 54.0 (10317) | 52.0 (251) | 56.0 (98) |
| pregnancy | 1.0 (31) | 0.0 (34) | 0.0 (47) | 1.0 (122) | 0.0 (2) | 0 (0) |
| **allergy** | | | | | | |
| allergy | 32.0 (1809) | 46.0 (5047) | 44.0 (4214) | 41.0 (7914) | 5.0 (23) | 39.0 (68) |
| no allergy | 68.0 (3769) | 54.0 (5925) | 56.0 (5394) | 59.0 (11273) | 95.0 (457) | 61.0 (106) |
| **smoking** | | | | | | |
| not smoking | 84.0 (4702) | 94.0 (10287) | 90.0 (8606) | 77.0 (14740) | 90.0 (430) | 81.0 (141) |
| smoking | 16.0 (876) | 6.0 (685) | 10.0 (1002) | 23.0 (4447) | 10.0 (50) | 19.0 (33) |

| | | | | | | |
|---|---|---|---|---|---|---|
| **self-assessment of cause** | | | | | | |
| cold | 20.0 (1101) | 35.0 (3822) | 15.0 (1407) | 27.0 (5259) | 17.0 (80) | 44.0 (76) |
| covid | 3.0 (153) | 9.0 (936) | 41.0 (3898) | 6.0 (1207) | 50.0 (240) | 13.0 (22) |
| flu | 62.0 (3432) | 38.0 (4192) | 17.0 (1588) | 29.0 (5567) | 14.0 (66) | 29.0 (51) |
| other | 16.0 (892) | 18.0 (2022) | 28.0 (2715) | 37.0 (7154) | 20.0 (94) | 14.0 (25) |
| **severity** | | | | | | |
| ILI fever | 40.0 (2206) | 15.0 (1689) | 20.0 (1912) | 33.0 (6412) | 23.0 (111) | 31.0 (54) |
| ILI fever phlegm | 12.0 (697) | 11.0 (1223) | 12.0 (1201) | 10.0 (1927) | 11.0 (54) | 5.0 (9) |
| ILI no fever | 48.0 (2675) | 73.0 (8060) | 68.0 (6495) | 57.0 (10848) | 66.0 (315) | 64.0 (111) |
| **period** | | | | | | |
| pandemic | 4.0 (250) | 23.0 (2528) | 100.0 (9608) | 15.0 (2918) | 100.0 (480) | 35.0 (61) |
| pre-pandemic | 92.0 (5143) | 77.0 (8444) | 0 (0) | 85.0 (16269) | 0 (0) | 65.0 (113) |

**Table S1. Descriptive statistics**. Unadjusted frequency of ILI episodes reported by InfluenzaNet individuals in Italy, the UK, the Netherlands, France, Belgium and Switzerland. Table shows for each country and category the percentage and occurrence of episodes and the fraction that occurred in the pre-pandemic and pandemic period. Individuals account for multiple ILI episodes across years. See Supplementary Tables S1-S2 for descriptive statistics specific to pre-pandemic and pandemic periods separately.

**Period-wise descriptive statistics**

| **variable** | **% of ILI episodes (occurrence)** | | | |
|---|---|---|---|---|
| **country** | **Italy** | **UK** | **France** | **Switzerland** |
| **medical service** | | | | |
| no | 61.97 (3187) | 77.36 (6532) | 54.72 (8903) | 80.53 (91) |
| yes | 38.03 (1956) | 22.64 (1912) | 45.28 (7366) | 19.47 (22) |
| **age class** | | | | |
| <15 | 9.62 (495) | 2.25 (190) | 7.24 (1178) | 8.85 (10) |
| 15-64 | 83.01 (4269) | 75.36 (6363) | 75.71 (12318) | 81.42 (92) |
| >65 | 7.37 (379) | 22.39 (1891) | 17.04 (2773) | 9.73 (11) |
| **gender** | | | | |
| Female | 48.61 (2500) | 70.78 (5977) | 68.47 (11140) | 70.8 (80) |
| Male | 51.39 (2643) | 29.22 (2467) | 31.53 (5129) | 29.2 (33) |
| **occupation** | | | | |
| not working | 43.81 (2253) | 38.76 (3273) | 46.27 (7527) | 27.43 (31) |
| working | 56.19 (2890) | 61.24 (5171) | 53.73 (8742) | 72.57 (82) |
| **education** | | | | |
| elementary | 23.72 (1220) | 10.0 (844) | 12.77 (2078) | 14.16 (16) |
| higher | 41.65 (2142) | 62.73 (5297) | 58.85 (9574) | 53.98 (61) |
| secondary | 34.63 (1781) | 27.27 (2303) | 28.38 (4617) | 31.86 (36) |

| | | | | |
|---|---|---|---|---|
| **duration** | | | | |
| <3 days | 4.67 (240) | 1.62 (137) | 3.15 (513) | 4.42 (5) |
| 3-6 days | 13.79 (709) | 4.89 (413) | 9.32 (1517) | 16.81 (19) |
| 7-10 days | 4.18 (215) | 2.16 (182) | 3.31 (538) | 6.19 (7) |
| >10 days | 3.31 (170) | 3.86 (326) | 2.97 (484) | 5.31 (6) |
| ND | 74.06 (3809) | 87.47 (7386) | 81.24 (13217) | 67.26 (76) |
| **co-morbidities** | | | | |
| yes | 19.41 (998) | 25.81 (2179) | 24.07 (3916) | 13.27 (15) |
| no | 80.59 (4145) | 74.19 (6265) | 75.93 (12353) | 86.73 (98) |
| **pregnancy** | | | | |
| NA | 59.62 (3066) | 44.1 (3724) | 45.21 (7355) | 40.71 (46) |
| no pregnancy | 39.8 (2047) | 55.5 (4686) | 54.13 (8807) | 59.29 (67) |
| pregnancy | 0.58 (30) | 0.4 (34) | 0.66 (107) | 0 (0) |
| **allergy** | | | | |
| allergy | 32.45 (1669) | 44.53 (3760) | 40.69 (6620) | 36.28 (41) |
| no allergy | 67.55 (3474) | 55.47 (4684) | 59.31 (9649) | 63.72 (72) |
| **smoking** | | | | |
| no smoking | 84.0 (4320) | 92.89 (7844) | 74.23 (12076) | 80.53 (91) |
| smoking | 16.0 (823) | 7.11 (600) | 25.77 (4193) | 19.47 (22) |
| **self-assessment of cause** | | | | |
| cold | 20.22 (1040) | 38.09 (3216) | 29.29 (4766) | 50.44 (57) |
| flu | 64.22 (3303) | 45.7 (3859) | 32.58 (5301) | 38.05 (43) |
| other | 15.56 (800) | 16.21 (1369) | 38.12 (6202) | 11.5 (13) |
| **severity** | | | | |
| ILI fever | 38.97 (2004) | 15.03 (1269) | 33.76 (5493) | 30.09 (34) |
| ILI fever phlegm | 12.66 (651) | 11.38 (961) | 9.98 (1624) | 4.42 (5) |
| ILI no fever | 48.38 (2488) | 73.59 (6214) | 56.25 (9152) | 65.49 (74) |

**Supplementary Table S2. Descriptive statistics of the pre-pandemic period**. Non-adjusted frequency of ILI episodes reported by InfluenzaNet individuals in Italy, the UK, the Netherlands, France, Belgium and Switzerland in the pre-pandemic period. Table shows for each country and category the percentage and occurrence of episodes. Individuals account for multiple ILI episodes across years.

| variable | % of ILI episodes (occurrence) | | | | | |
|---|---|---|---|---|---|---|
| **country** | **Italy** | **UK** | **Netherlands** | **France** | **Belgium** | **Switzerland** |
| **medical service** | | | | | | |
| no | 40.8 (102) | 78.92 (1995) | 82.69 (7945) | 58.12 (1696) | 60.21 (289) | 73.77 (45) |
| yes | 59.2 (148) | 21.08 (533) | 17.31 (1663) | 41.88 (1222) | 39.79 (191) | 26.23 (16) |
| **age class** | | | | | | |
| <15 | 5.2 (13) | 1.7 (43) | 0.87 (84) | 4.08 (119) | 2.71 (13) | 6.56 (4) |
| 15-64 | 84.4 (211) | 69.22 (1750) | 84.32 (8101) | 74.19 (2165) | 70.21 (337) | 75.41 (46) |
| >65 | 10.4 (26) | 29.07 (735) | 14.81 (1423) | 21.73 (634) | 27.08 (130) | 18.03 (11) |
| **gender** | | | | | | |
| Female | 54.4 (136) | 73.81 (1866) | 70.57 (6780) | 66.45 (1939) | 71.67 (344) | 63.93 (39) |
| Male | 45.6 (114) | 26.19 (662) | 29.43 (2828) | 33.55 (979) | 28.33 (136) | 36.07 (22) |
| **occupation** | | | | | | |
| not working | 38.4 (96) | 41.77 (1056) | 31.38 (3015) | 44.07 (1286) | 41.25 (198) | 39.34 (24) |
| working | 61.6 (154) | 58.23 (1472) | 68.62 (6593) | 55.93 (1632) | 58.75 (282) | 60.66 (37) |
| **education** | | | | | | |
| elementary | 6.8 (17) | 22.15 (560) | 3.31 (318) | 5.79 (169) | 5.42 (26) | 11.48 (7) |
| higher | 44.8 (112) | 58.82 (1487) | 58.03 (5576) | 69.84 (2038) | 72.5 (348) | 68.85 (42) |
| secondary | 48.4 (121) | 19.03 (481) | 38.66 (3714) | 24.37 (711) | 22.08 (106) | 19.67 (12) |
| **duration** | | | | | | |
| <3 days | 8.4 (21) | 4.15 (105) | 8.24 (792) | 4.22 (123) | 5.62 (27) | 6.56 (4) |
| 3-6 days | 13.2 (33) | 6.17 (156) | 13.0 (1249) | 10.32 (301) | 8.75 (42) | 16.39 (10) |
| 7-10 days | 2.4 (6) | 3.36 (85) | 4.28 (411) | 3.84 (112) | 2.08 (10) | 9.84 (6) |
| >10 days | 3.6 (9) | 3.56 (90) | 2.54 (244) | 3.56 (104) | 1.04 (5) | 4.92 (3) |
| ND | 72.4 (181) | 82.75 (2092) | 71.94 (6912) | 78.07 (2278) | 82.5 (396) | 62.3 (38) |
| **co-morbidities** | | | | | | |
| yes | 21.6 (54) | 31.33 (792) | 25.75 (2474) | 28.44 (830) | 54.17 (260) | 11.48 (7) |
| no | 78.4 (196) | 68.67 (1736) | 74.25 (7134) | 71.56 (2088) | 45.83 (220) | 88.52 (54) |
| **pregnancy** | | | | | | |
| NA | 50.0 (125) | 47.86 (1210) | 37.83 (3635) | 47.74 (1393) | 47.29 (227) | 49.18 (30) |
| no pregnancy | 50.0 (125) | 52.14 (1318) | 61.68 (5926) | 51.75 (1510) | 52.29 (251) | 50.82 (31) |
| pregnancy | 0 (0) | 0 (0) | 0.49 (47) | 0.51 (15) | 0.42 (2) | 0 (0) |
| **allergy** | | | | | | |
| allergy | 36.0 (90) | 50.91 (1287) | 43.86 (4214) | 44.35 (1294) | 4.79 (23) | 44.26 (27) |
| no allergy | 64.0 (160) | 49.09 (1241) | 56.14 (5394) | 55.65 (1624) | 95.21 (457) | 55.74 (34) |
| **smoking** | | | | | | |
| not smoking | 88.8 (222) | 96.64 (2443) | 89.57 (8606) | 91.3 (2664) | 89.58 (430) | 81.97 (50) |
| smoking | 11.2 (28) | 3.36 (85) | 10.43 (1002) | 8.7 (254) | 10.42 (50) | 18.03 (11) |
| **self-assessment of cause** | | | | | | |
| cold | 18.8 (47) | 23.97 (606) | 14.64 (1407) | 16.9 (493) | 16.67 (80) | 31.15 (19) |

| | | | | | | |
|---|---|---|---|---|---|---|
| covid | 42.8 (107) | 37.03 (936) | 40.57 (3898) | 41.36 (1207) | 50.0 (240) | 36.07 (22) |
| flu | 19.2 (48) | 13.17 (333) | 16.53 (1588) | 9.12 (266) | 13.75 (66) | 13.11 (8) |
| other | 19.2 (48) | 25.83 (653) | 28.26 (2715) | 32.63 (952) | 19.58 (94) | 19.67 (12) |

**Supplementary Table S3. Descriptive statistics of the pandemic period**. Non-adjusted frequency of ILI episodes reported by InfluenzaNet individuals in Italy, the UK, the Netherlands, France, Belgium and Switzerland. Table shows for each country and category the percentage and occurrence of episodes and if they occurred in the pre-pandemic or pandemic period. Individuals account for multiple ILI episodes across years.

### Sensitivity analysis on ILI case definition

We replicate the analysis using the WHO case definition and compare adjusted odds-ratios and unadjusted rates of HCSB for Italy and France.

| **variable** | **ORc (95%CI)** | | | |
|---|---|---|---|---|
| **country** | **Italy** | **Italy (WHO)** | **France** | **France (WHO)** |
| **age** | | | | |
| 15-64 | | | | |
| <15 | **1.62 (1.44-1.8)** | **1.44 (1.15-1.72)** | **1.33 (1.22-1.44)** | **1.22 (1.06-1.36)** |
| >65 | **1.34 (1.15-1.54)** | 0.97 (0.69-1.28) | **1.15 (1.06-1.24)** | 1.08 (0.95-1.21) |
| **comorbidities** | | | | |
| no | | | | |
| yes | **1.29 (1.18-1.4)** | **1.39 (1.2-1.58)** | **1.21 (1.16-1.26)** | **1.19 (1.11-1.26)** |
| **pregnancy** | | | | |
| no pregnancy | | | | |
| pregnancy | 0.89 (0.46-1.46) | 1.04 (0.35-2.0) | **1.34 (1.05-1.61)** | 1.29 (0.77-1.64) |
| NA | **0.8 (0.66-0.96)** | 0.89 (0.66-1.17) | 0.96 (0.88-1.04) | 0.94 (0.83-1.05) |
| **smoking** | | | | |
| not smoking | | | | |
| smoking | 1.08 (0.97-1.19) | 1.03 (0.84-1.22) | 0.96 (0.91-1.0) | 0.94 (0.86-1.01) |
| **allergy status** | | | | |
| no allergy | | | | |
| allergy | 0.95 (0.87-1.04) | 0.92 (0.78-1.07) | 1.01 (0.97-1.06) | 0.94 (0.87-1.0) |
| **severity** | | | | |
| ILI no fever | | | | |
| ILI fever | **1.76 (1.61-1.9)** | **-** | **1.55 (1.49-1.6)** | - |
| ILI fever phlegm | **2.37 (2.16-2.57)** | **1.34 (1.18-1.5)** | **1.75 (1.66-1.83)** | **1.15 (1.07-1.22)** |
| **gender** | | | | |

| | | | | |
|---|---|---|---|---|
| Male | | | | |
| Female | 0.99 (0.85-1.13) | 0.87 (0.69-1.06) | 1.0 (0.94-1.07) | 0.95 (0.87-1.04) |
| **occupation** | | | | |
| not working | | | | |
| working | **1.19 (1.08-1.3)** | 1.08 (0.89-1.27) | 1.04 (0.99-1.09) | 1.08 (1.0-1.16) |
| **education** | | | | |
| secondary | | | | |
| elementary | 1.1 (0.96-1.23) | 0.99 (0.77-1.21) | 0.92 (0.84-1.0) | 1.03 (0.9-1.16) |
| higher | 0.95 (0.86-1.05) | 0.85 (0.7-1.0) | **0.84 (0.8-0.88)** | **0.9 (0.83-0.98)** |
| **self-assessment of cause** | | | | |
| cold | | | | |
| flu | **2.81 (2.45-3.19)** | **2.03 (1.54-2.57)** | **2.0 (1.9-2.11)** | **1.79 (1.63-1.95)** |
| other | **3.24 (2.8-3.7)** | **2.49 (1.83-3.16)** | **2.85 (2.75-2.95)** | **2.86 (2.73-2.97)** |
| **duration** | | | | |
| <3 days | | | | |
| 3-6 days | **2.4 (1.9-2.93)** | **1.87 (1.19-2.66)** | **2.27 (1.96-2.6)** | **1.63 (1.33-1.91)** |
| 7-10 days | **3.24 (2.59-3.84)** | **2.3 (1.41-3.24)** | **3.1 (2.7-3.5)** | **2.02 (1.7-2.29)** |
| >10 days | **3.48 (2.83-4.07)** | **2.16 (1.06-3.4)** | **4.13 (3.76-4.46)** | **1.82 (1.32-2.26)** |
| ND | **2.53 (2.06-3.02)** | **2.01 (1.33-2.77)** | **2.98 (2.67-3.29)** | **1.79 (1.52-2.04)** |

**Supplementary Table S4. Pre-pandemic determinants of HCSB.** Table shows for Italy and France the adjusted odds-ratios of healthcare seeking behavior and the 95% confidence intervals in parentheses based on the ECDC and the WHO ILI case definitions respectively. Reference groups for each covariate are reported at the first row. Odds-ratios are adjusted by the Zhang-Yu correction. Odds-ratios for some groups are not computed due to absence of case occurrence in the class. †Group sample lower than 5%. ND = No Data. Severity odds-ratios for WHO case definition were computed with respect to the WHO ILI fever category, as fever is a requirement for WHO definition.

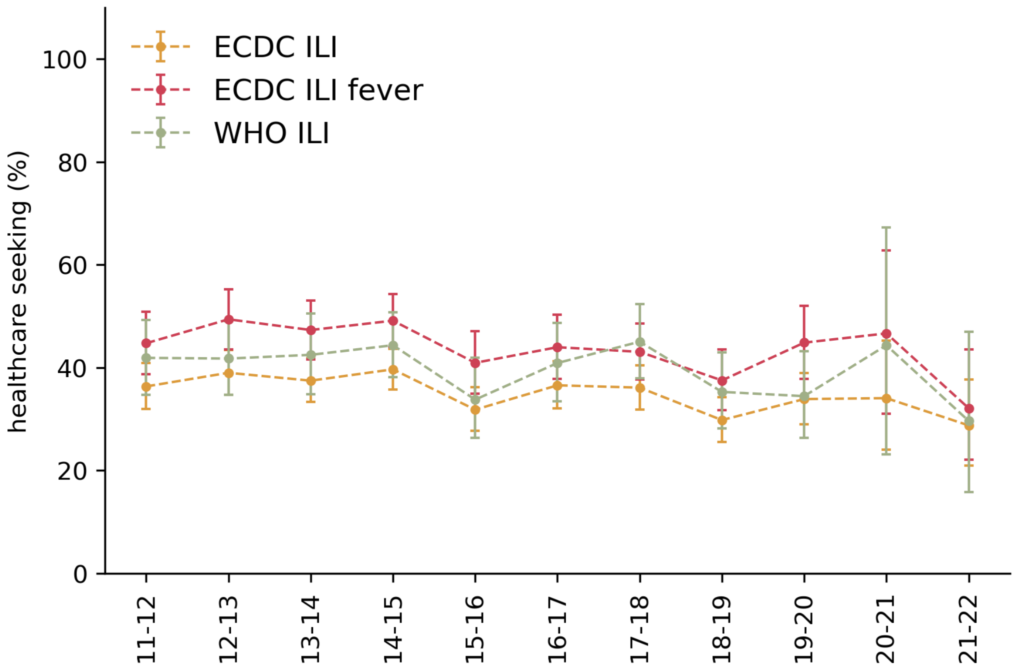


**Supplementary Figure S2. WHO and ECDC case definitions comparison of healthcare seeking behavior for ILI patients in Italy.** Comparison of healthcare seeking behavior from online cohorts in Italy among ECDC and WHO ILI case definitions. *ECDC ILI* and *WHO ILI* account for all episodes matching ECDC and WHO definitions respectively. *ECDC ILI* is a subset of *ECDC ILI* episodes with fever shown for comparison with the WHO case definition requiring fever. Whiskers represent the 95% confidence interval computed through the Clopper-Pearson method.

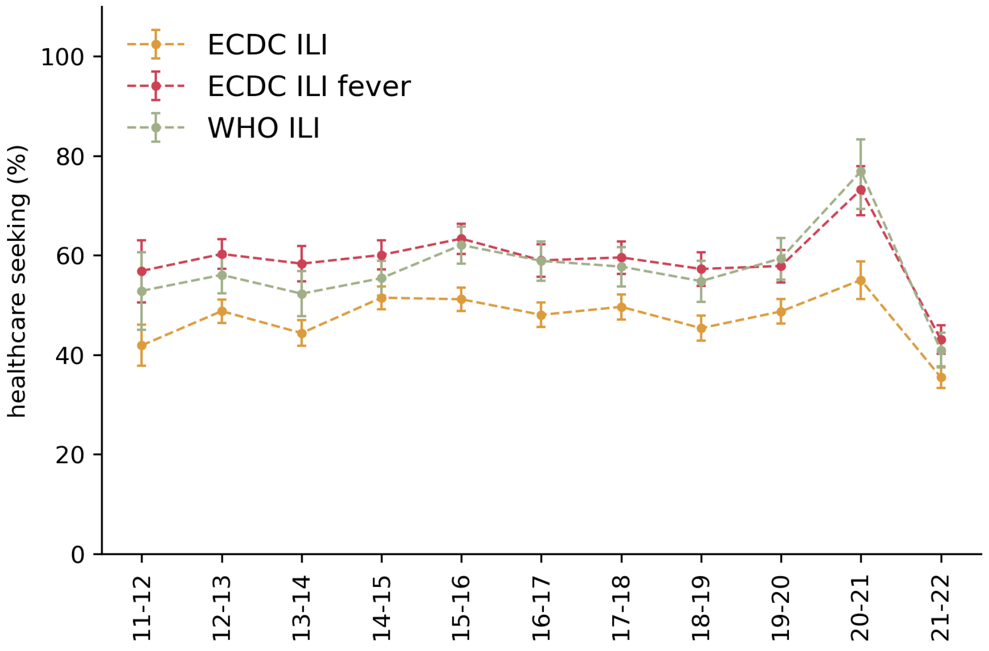


**Supplementary Figure S3. WHO and ECDC case definitions comparison of healthcare seeking behavior for ILI patients in France.** Comparison of healthcare seeking behavior from online cohorts in Italy among ECDC and WHO ILI case definitions. *ECDC ILI* and *WHO ILI* account for all episodes matching ECDC and WHO definitions respectively. *ECDC ILI* is a subset of *ECDC ILI* episodes with fever shown for comparison with the WHO case definition requiring fever. Whiskers represent the 95% confidence interval computed through the Clopper-Pearson method.

**ILI case definition**

We report the ILI case definitions employed in the past and nowadays by national surveillance networks and the ECDC and WHO proposed ILI case definitions in **Table S1**.

| **Surveillance System** | **past ILI case definition** | **new ILI case definition** | **source** |
|---|---|---|---|
| **Italy (Istituto Superiore di Sanità)** | Acute onset of fever > 38°C<br>AND<br>respiratory symptoms<br>AND<br>one of these symptoms:<br>- headache<br>- general discomfort<br>- asthenia | ECDC definition | 1,2 |
| **France (Réseau Sentinelles)** | Sudden onset of fever (>39 °C) AND respiratory symptoms<br>AND<br>stiffness, myalgia | ceased surveillance on ILI, now reporting ARI:<br>Sudden onset<br>AND<br>fever<br>AND<br>respiratory symptoms | 3,4 |
| **UK (UKHSA Royal College of GPs Sentinel Network, England)** | Acute respiratory infection<br>AND<br>fever of ≥ 38 C° (measured or plausibly)<br>AND<br>cough<br>AND<br>onset within the last 10 days<br>AND<br>Sudden onset<br>OFTEN<br>symptoms of systems upset like myalgia, fatigue, malaise, headache, etc.<br>WITHOUT<br>another more plausible diagnosis.<br>"RCGP RSC stresses the notion of symptoms within 10 days of onset to differentiate acute episodes, with swabs only wanted within 7 days of onset." | Acute respiratory infection<br>AND<br>fever of ≥ 38 C° (measured or plausibly)<br>AND<br>cough<br>AND<br>onset within the last 10 days<br>AND<br>Sudden onset<br>OFTEN<br>symptoms of systems upset like myalgia, fatigue, malaise, headache, etc.<br>WITHOUT<br>another more plausible diagnosis.<br>"RCGP RSC stresses the notion of symptoms within 10 days of onset to differentiate acute episodes, with swabs only wanted within 7 days of onset." | 3,5 |
| **Netherlands (Nivel Primary Care)** | Pel-criteria:<br>sudden onset of symptoms<br>AND<br>fever (at least 38 °C, rectal temperature)<br>AND<br>at least one of the following:<br>- cough<br>- rhinorrhoea<br>- sore throat<br>- frontal headache<br>- retrosternal pain<br>- myalgia | Pel-criteria:<br>sudden onset of symptoms<br>AND<br>fever (at least 38 °C, rectal temperature)<br>AND<br>at least one of the following:<br>- cough<br>- rhinorrhoea<br>- sore throat<br>- frontal headache<br>- retrosternal pain<br>- myalgia | 3,6 |
| **Belgium (Sciensano)** | sudden onset with fever<br>AND<br>myalgia<br>AND<br>respiratory symptoms (cough or thoracic pain) | WHO definition<br>AND<br>sudden onset of symptoms | 3,7 |
| **Switzerland (Sentinella)** | Respiratory illness | sudden onset of high fever (>38°C) | 3,8 |

| | | | |
|---|---|---|---|
| | AND<br>fever > 38 °C<br>AND<br>myalgia<br>AND<br>general pain<br>AND<br>chills<br>AND<br>anorexia (optional symptoms are: cough, rhinitis and arthralgia) | AND<br>cough OR sore throat | |
| **ECDC** | | Sudden onset of symptoms<br>AND<br>at least one of the following:<br>- Fever or feverishness<br>- Malaise<br>- Headache<br>- Myalgia<br>AND<br>at least one of the following:<br>- Cough<br>- Sore throat<br>- Shortness of breath | 9 |
| **WHO** | | Acute respiratory infection<br>AND<br>fever of ≥ 38 C°<br>AND<br>cough<br>AND<br>onset within the last 10 days | 10 |

**Table S5: ILI definitions adopted by the national surveillance systems.** ILI case definitions adopted in the pre-H1N1 pandemic by national surveillance systems and ILI definitions adopted nowadays.

**Previous estimates of ILI burden in Italy**

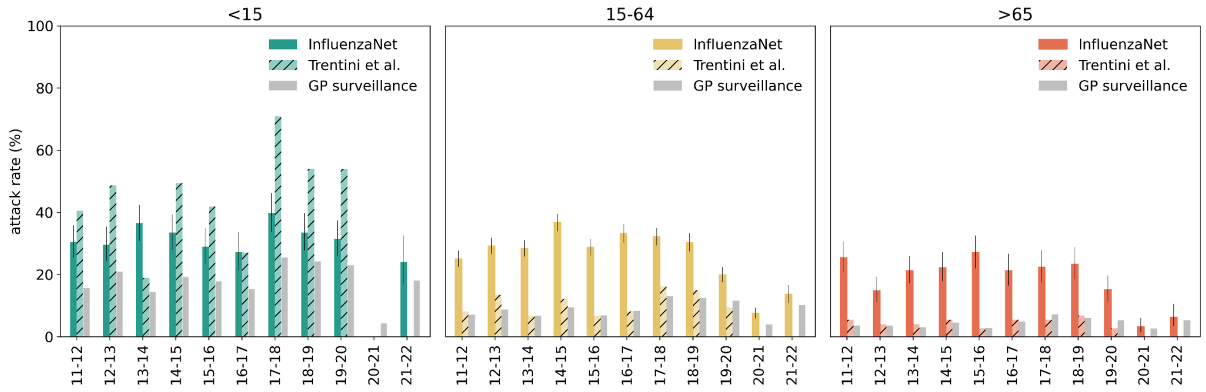


**Supplementary Figure S4. Comparison of ILI burden of cases for Italy.** National comparison of InfluenzaNet reported ILI burden, InfluenzaNet as colored bars, estimates by Trentini et al.[1] as hatched bars and GP surveillance reported burden as gray bars, across years. Attack rates are expressed in percentage. Whiskers representing the 95%CIs.

**References**


1. Trentini F, Pariani E, Bella A, et al. Characterizing the transmission patterns of seasonal influenza in Italy: lessons from the last decade. *BMC Public Health*. 2022;22(1):19. doi:10.1186/s12889-021-12426-9

2. EpiCentro, Istituto Superiore di Sanità. Definizione di caso. Accessed March 4, 2024. https://www.epicentro.iss.it/influenza/definizione-caso

3. Aguilera JF, Paget WJ, Mosnier A, et al. Heterogeneous case definitions used for the surveillance of influenza in Europe. *Eur J Epidemiol*. 2003;18(8):751-754. doi:10.1023/A:1025337616327

4. Coletti P, Poletto C, Turbelin C, Blanchon T, Colizza V. Shifting patterns of seasonal influenza epidemics. *Sci Rep*. 2018;8(1):12786. doi:10.1038/s41598-018-30949-x

5. Elson WH, Jamie G, Wimalaratna R, et al. Validation of an acute respiratory infection phenotyping algorithm to support robust computerised medical record-based respiratory sentinel surveillance, England, 2023. *Eurosurveillance*. 2024;29(35):2300682. doi:10.2807/1560-7917.ES.2024.29.35.2300682

6. Reukers DFM, Van Asten L, Brandsema PS, et al. Annual report Surveillance of influenza and other respiratory infections: Winter 2017/2018. Published online 2018. doi:10.21945/RIVM-2018-0049

7. Sciensano. Influenza and influenza-like illness. For a Healthy Belgium. September 6, 2020. Accessed March 4, 2024. https://www.healthybelgium.be/en/health-status/communicable-diseases/influenza-and-influenza-like-illness

8. Federal Office of Public Health Infectious Diseases Dashboard (IDD). Accessed March 4, 2024. https://idd.bag.admin.ch/

9. EU case definitions. June 7, 2018. Accessed February 12, 2024. https://www.ecdc.europa.eu/en/all-topics/eu-case-definitions

10. Casalegno JS, Eibach D, Valette M, et al. Performance of influenza case definitions for influenza community surveillance: based on the French influenza surveillance network GROG, 2009-2014. *Eurosurveillance*. 2017;22(14):30504. doi:10.2807/1560-7917.ES.2017.22.14.30504
